# Deep Learning-powered CT-less Multi-tracer Organ Segmentation from PET Images: A solution for unreliable CT segmentation in PET/CT Imaging

**DOI:** 10.1101/2024.08.27.24312482

**Authors:** Yazdan Salimi, Zahra Mansouri, Isaac Shiri, Ismini Mainta, Habib Zaidi

## Abstract

**Introduction:** The common approach for organ segmentation in hybrid imaging relies on co-registered CT (CTAC) images. This method, however, presents several limitations in real clinical workflows where mismatch between PET and CT images are very common. Moreover, low-dose CTAC images have poor quality, thus challenging the segmentation task. Recent advances in CT-less PET imaging further highlight the necessity for an effective PET organ segmentation pipeline that does not rely on CT images. Therefore, the goal of this study was to develop a CT-less multi-tracer PET segmentation framework.

**Methods:** We collected 2062 PET/CT images from multiple scanners. The patients were injected with either ^18^F-FDG (1487) or ^68^Ga-PSMA (575). PET/CT images with any kind of mismatch between PET and CT images were detected through visual assessment and excluded from our study. Multiple organs were delineated on CT components using previously trained in-house developed nnU-Net models. The segmentation masks were resampled to co-registered PET images and used to train four different deep-learning models using different images as input, including non-corrected PET (PET-NC) and attenuation and scatter-corrected PET (PET-ASC) for ^18^F-FDG (tasks #1 and #2, respectively using 22 organs) and PET-NC and PET-ASC for ^68^Ga tracers (tasks #3 and #4, respectively, using 15 organs). The models’ performance was evaluated in terms of Dice coefficient, Jaccard index, and segment volume difference.

**Results:** The average Dice coefficient over all organs was 0.81±0.15, 0.82±0.14, 0.77±0.17, and 0.79±0.16 for tasks #1, #2, #3, and #4, respectively. PET-ASC models outperformed PET-NC models (P-value < 0.05). The highest Dice values were achieved for the brain (0.93 to 0.96 in all four tasks), whereas the lowest values were achieved for small organs, such as the adrenal glands. The trained models showed robust performance on dynamic noisy images as well.

**Conclusion:** Deep learning models allow high performance multi-organ segmentation for two popular PET tracers without the use of CT information. These models may tackle the limitations of using CT segmentation in PET/CT image quantification, kinetic modeling, radiomics analysis, dosimetry, or any other tasks that require organ segmentation masks.

## Introduction

PET/CT hybrid imaging provides valuable information by combining structural, molecular, and physiological information with a wide range of indications and radiopharmaceuticals [1]. Since the emergence of hybrid PET/CT imaging, the development and use of different radiotracers has expanded increased. ^18^F-Fluorodeoxyglucose (^18^F-FDG), with a wide range of indications for brain, cardiac, and oncological imaging, is the most common used radiotracer in clinical practice [2–4]. Other common radiotracers are prostate-specific membrane antigen radiolabeled ligands, such as ^68^Ga-PSMA and radiolabeled somatostatin analogues, such as ^68^Ga-DOTATATE. ^68^Ga-PSMA has high diagnostic accuracy in the initial staging and biochemical recurrence evaluation in patients diagnosed with prostate cancer [5; 6]. It is also used in the context of PSMA radioligand theragnostics, for lesions’ evaluation and patient selection with a good predictive value [7].

Medical image segmentation in general and in nuclear medicine in particular, is a crucial step towards modern personalized medicine. Segmentation can play an important role in PET image quantification in each organ and volume of interest (VOI) [8; 9]. Quantitative PET provides detailed information for accurate diagnosis and therapy by precise measurement of tracer uptakes and kinetics within each VOI [10; 11]. Nowadays, with the growing interest in personalized dosimetry for radiopharmaceutical therapy (RPT), aiming at delivering a tumoricidal radiation dose to the target volume while sparing organs at risk (OARs) [12; 13], the importance of image segmentation is becoming more pronounced. Moreover, the assessment of treatment response and prognostication, which requires tumor and OAR masks, have recently received significant attention [14–18]. In addition, studies highlighted the importance of non-tumoral organs in prognostication and outcome prediction [19; 20].

In clinical practice, segmentation is performed manually on CT or MRI images after visual inspection of PET images [21–23]. Manual contouring is subjective, time-consuming, labor intensive, and prone to errors and inter-/intra-operator variability because of different levels of expertise and the use of different windowing settings [24; 25]. The available methods for automated organ segmentation in hybrid molecular imaging, predominantly using deep learning (DL), focus on using co-registered CT images [26]. Reliable CT segmentation tools capable of automated segmentation [27; 28] can be used for PET/CT images. However, this approach faces three main limitations.

First, mismatch between emission (PET/SPECT) and transmission (CT) images is highly prevalent in clinical setting [29; 30]. This issue becomes more challenging in dynamic imaging protocols, where CT images are acquired within seconds at the beginning of the exam, while the dynamic PET scan is usually acquired during much longer time, including inevitably averaging multiple respiratory and cardiac cycles. In addition, involuntary changes in the position and size of the organs, such as the bladder getting filled and bowel movements, limit CT segmentation reliability [31]. Additionally, patient bulk motion during prolonged dynamic acquisitions further complicate alignment. Second, the low-dose and ultra-low-dose attenuation correction CT (CTAC) images acquired using lower tube currents and special beam filtering [32], often used in PET/CT, suffer from reduced image quality, thus affecting the accuracy of segmentation. CTAC. Last, the potential advent of CT-less clinical scanners, such as PET-only and PET/MRI scanners [30; 33], which utilize DL-based or Maximum Likelihood estimation of Activity and Attenuation (MLAA)-based attenuation correction methods, pose a significant challenge to CT-based segmentation approaches. DL-guided MRI multi-organ segmentation models were recently introduced to overcome PET/MRI organ segmentation [34]. These limitations highlight the necessity for developing segmentation tools based on emission data only rather than relying on co-registered CT images.

Utilizing the emission data to improve the performance of DL-based organ segmentation has been previously reported [35–38]. Klyuzhin *et al.* [35] used both PET and CT images for improved organ segmentation in PET/CT. Yazdani *et al.* [36] developed DL segmentation models to segment both healthy organs and malignant lesions from ^68^Ga-PSMA PET/CT images and compared them to using only PET, only CT, and both images as input to their DL models. Wang *et al.* [37] segmented bladder and heart on ^18^F-FDG PET/CT images to overcome the issue of absence or availability of unreliable CT images. Clement *et al.* [38] developed a model to perform CT-less organ segmentation on ^18^F-FDG PET images for dynamic imaging.

This study aimed to develop a reliable CT-less multi-organ segmentation pipeline on two common radiotracers (^18^F-FDG and ^68^Ga-PSMA) using a multi-centric dataset to address the limitations of CT-based segmentation approaches in hybrid PET/CT imaging.

## Materials and Methods

### Common processing and steps for both tracers for reference segmentation generation

Three types of images, including CTAC, Non-corrected PET (NC), and attenuation and scatter-corrected (CT-ASC) PET images were collected in a fully anonymized setup. All images were visualized using the open-source ITK-SNAP software [39]. Images presenting with a mismatch between PET and CT were excluded from training whereas PET/CT images without mismatch were included in the next steps. Figure 1 illustrates an example of a mismatch visualized with segmentation generated based on the co-registered CT scan. Using previously developed DL-based segmentation models in our group [28] based on nnU-Net architecture [40], a total number of 22 organs were delineated on the CTAC component of PET/CT images. All nnU-Net five folds were assembled on images to ensure the highest segmentation accuracy. The previously trained models were separate models, each dedicated to one specific organ. The CT-generated segmentation masks were dilated by 2 mm and resampled with the co-registered PET image voxel spacing. During down sampling from CT voxel spacing (1 to 1.5 mm) to PET spacing (1.6 to 4 mm) without dilation, certain organ shapes, such as ribs and thin parts of pelvic hip bones, may be lost. This occurs because nearest neighborhood interpolation, which is necessary for maintaining a binary segment with values of 0 and 1, has limitations that can result in the removal of fine details in the segmentation. The segmentations were integrated into a unified multi-value segmentation mask using the Simple ITK 2.2 Python library, prioritizing organs with higher Dice values from CT training task. For instance, if a single voxel was segmented as both liver and stomach by separate CT segmentation models, the voxel was classified as liver tissue to avoid overlap between the two organs. The segmentation masks and PET images were used to train an nnU-Net model using a five-fold cross-validation data split. Four different tasks were defined: two utilizing ^18^F-FDG PET images and two utilizing ^68^Ga-PET images. Each task involved using either ASC or NC as inputs, namely, ^18^F-FDG-NC (task #1), ^18^F-FDG-ASC (task #2), ^68^Ga-NC (task #3), and ^68^Ga-ASC (task #4).

**Figure 1.**
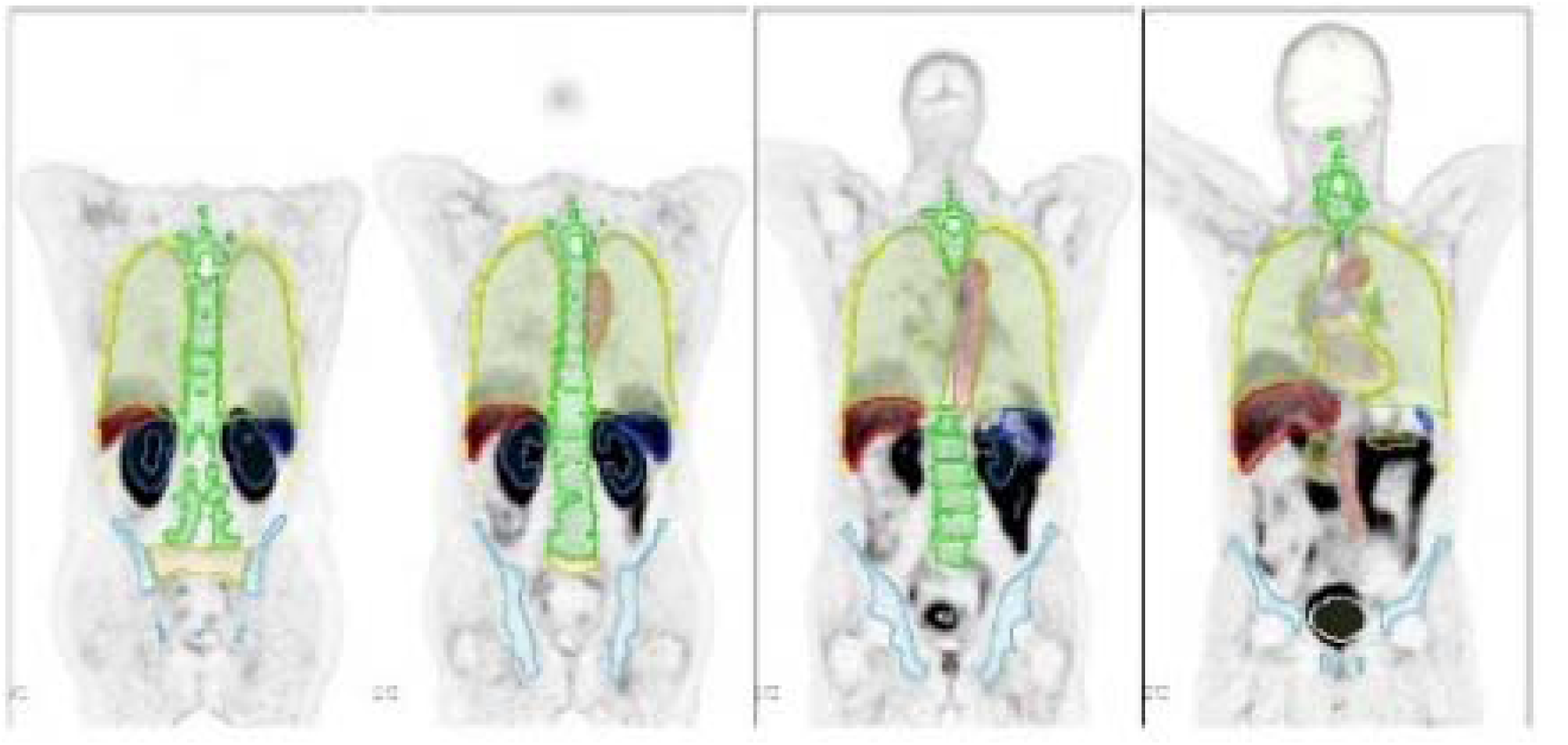
Coronal slices showing representative cases presenting with mismatch between CT-based segmentation and PET images. Please note the chest/abdomen interval organs.

**Figure 2.**
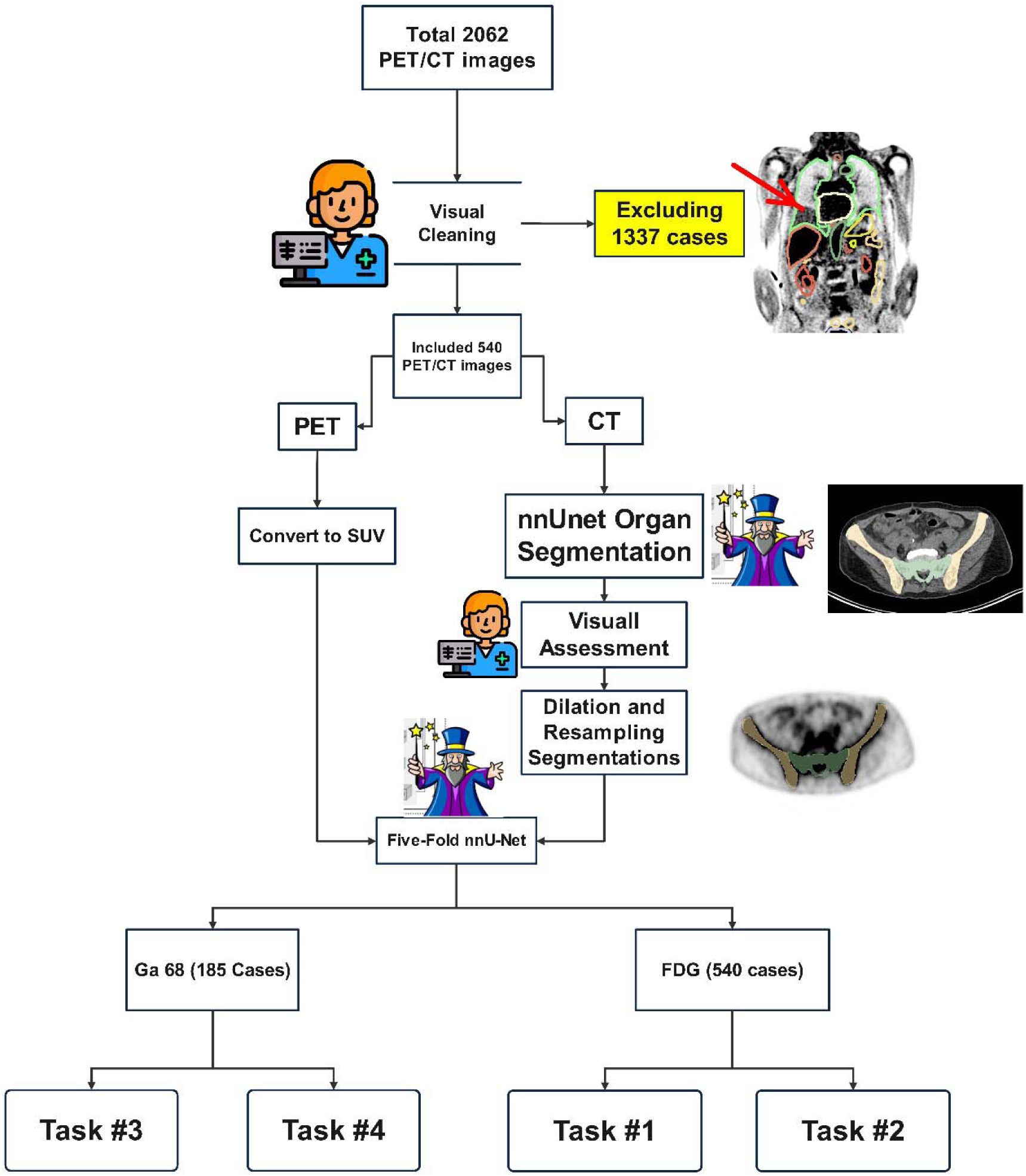
Flowchart of steps followed in this study, including generation of reference segmentation masks and training the LD-nnU-Net models.

### Datasets

This study included two separate sets of images acquired from patients injected with ^18^F-FDG (dataset #1) and ^68^Ga-PSMA tracers (dataset#2). The study was approved by the local ethics committee and consent was waived owing to the retrospective nature of the study protocol.

*Dataset #1 (^18^F-FDG).* This dataset included patients injected with ^18^F-FDG for oncological indications, with whole-body PET/CT images acquired on Biograph mCT and Biograph Vision scanners (Siemens Healthineers, TN, USA). Initially, 1487 images were included. After excluding PET/CT image pairs with mismatches, that is 947 image pairs (∼64%), 540 cases remained for five-fold training. The semi-diagnostic CTAC images acquired with an average tube current of ∼110 mAs, and PET-NC and PET-ASC images were reconstructed using iterative reconstruction methods. Detailed information about dataset #1 can be found in Table 1. A total number of 22 organs were selected for tasks #1 and #2 on dataset #1, including the adrenal gland (AG), Aorta, Colon, Esophagus, Eyeballs, Femoral Heads (FH), Gall bladder (GB), Heart, Hip bones (including Ilium, Ischium, and Pubis as a single mask), Kidneys, Liver, Lungs, Pancreas, Erectus Spinae, Rib Cage, Sacrum, Spleen, Stomach, Urinary bladder (UB), Vertebrae, Brain, and Clavicle.

**Table 1.**
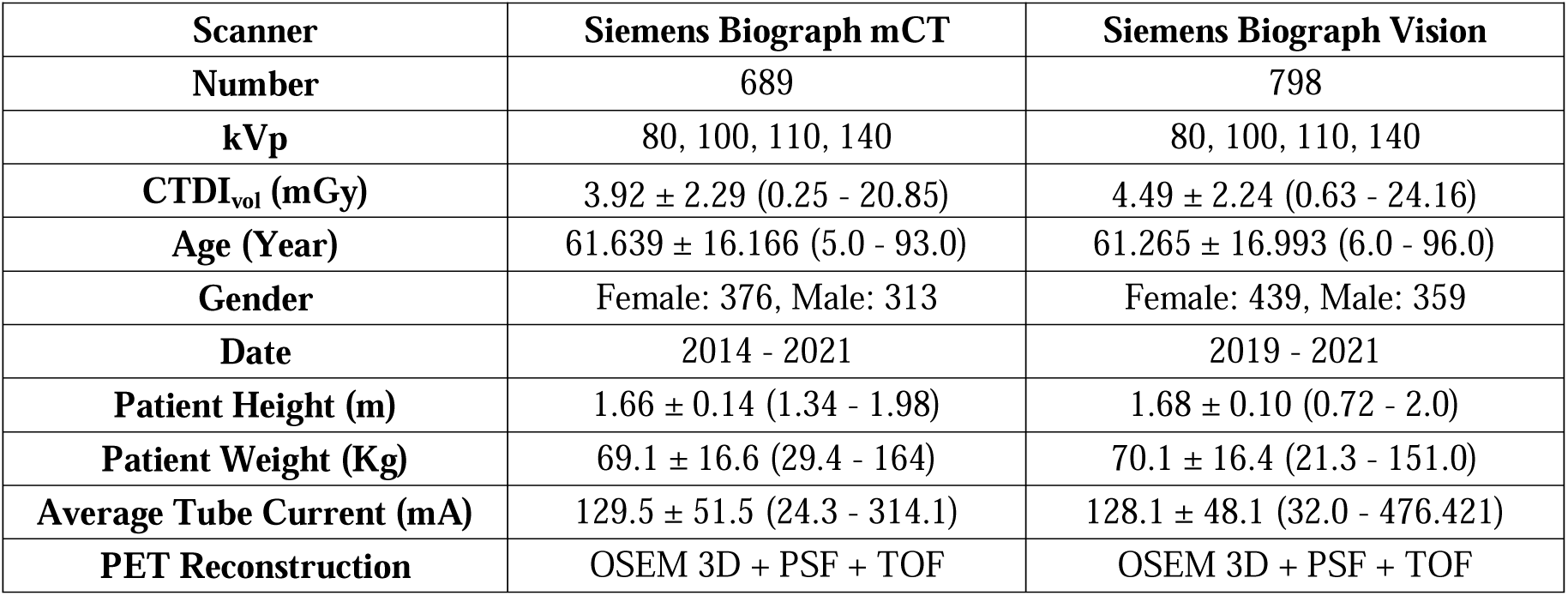
Detailed demographics of all 1487 images included as dataset #1 (^18^F-FDG).

From the 575 cases initially collected, 390 were excluded, leaving only 185 clean cases for the remainder of our study. Table 2 summarizes the demographic information for all 575 cases initially included in our study. Some information was missing due to the anonymization process. A total number of 15 organs were selected for tasks #3 and #4 on dataset #2, including AG aorta, Brain, Eyeballs, Hip bones, Kidneys, Liver, Lungs, Pancreas, Rib cage, Sacrum, Spleen, UB, Vertebrae, Heart. Seven organs were excluded from dataset #2 for tasks #3 and #4 as there is less anatomical information in ^68^Ga images compared to ^18^F-FDG images. We aimed to include only organs with distinguishable uptake.

**Table 2.**
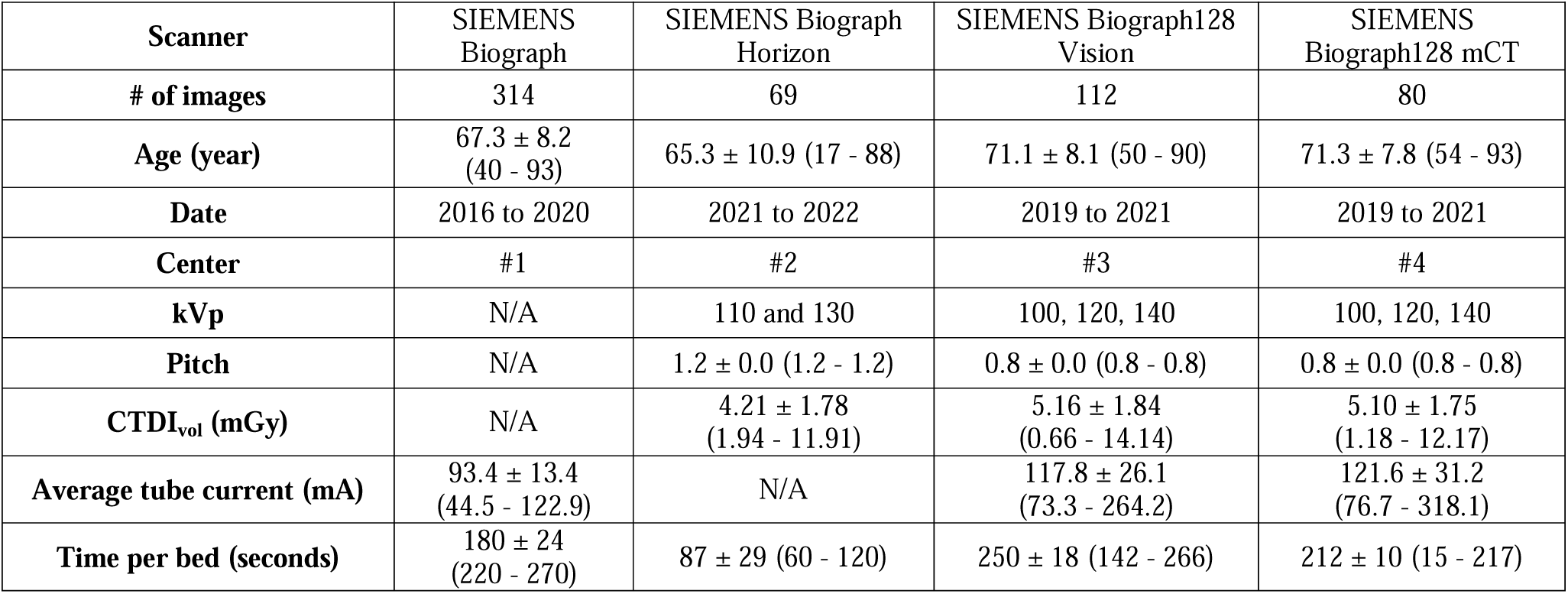
Demographic information of all included 575 ^68^Ga-PET/CT images from dataset #2 in this study.

| Scanner | SIEMENS Biograph | SIEMENS Biograph Horizon | SIEMENS Biograph128 Vision | SIEMENS Biograph128 mCT |
| --- | --- | --- | --- | --- |
| # of images | 314 | 69 | 112 | 80 |
| Age (year) | $67.3 \pm 8.2$<br>(40 - 93) | $65.3 \pm 10.9$ (17 - 88) | $71.1 \pm 8.1$ (50 - 90) | $71.3 \pm 7.8$ (54 - 93) |
| Date | 2016 to 2020 | 2021 to 2022 | 2019 to 2021 | 2019 to 2021 |
| Center | #1 | #2 | #3 | #4 |
| kVp | N/A | 110 and 130 | 100, 120, 140 | 100, 120, 140 |
| Pitch | N/A | $1.2 \pm 0.0$ (1.2 - 1.2) | $0.8 \pm 0.0$ (0.8 - 0.8) | $0.8 \pm 0.0$ (0.8 - 0.8) |
| CTDI<sub>vol</sub> (mGy) | N/A | $4.21 \pm 1.78$<br>(1.94 - 11.91) | $5.16 \pm 1.84$<br>(0.66 - 14.14) | $5.10 \pm 1.75$<br>(1.18 - 12.17) |
| Average tube current (mA) | $93.4 \pm 13.4$<br>(44.5 - 122.9) | N/A | $117.8 \pm 26.1$<br>(73.3 - 264.2) | $121.6 \pm 31.2$<br>(76.7 - 318.1) |
| Time per bed (seconds) | $180 \pm 24$<br>(220 - 270) | $87 \pm 29$ (60 - 120) | $250 \pm 18$ (142 - 266) | $212 \pm 10$ (15 - 217) |

### Model training parameters

Four separated nnU-Net [40] models were trained for the four defined tasks. For each task, the combined segmentation masks and the corresponding PET images were fed into an nnU-Net version 2 (nnunetv2) pipeline using default parameters except the training length which was increased from the default value of 1000 epochs to 2000 epochs to enhance accuracy. We utilized nnU-Net 3D-fullres training configuration, which uses 3D patches for training. The initial learning rate was set to 1e-2 and decreased every epoch. The decay of 3e-5 and the Dice cross-entropy loss function were used. Five-fold cross-validation data splits were used, with 80% of images used for training and 20% for testing in each fold. The training process was conducted on a PC equipped with an RTX4090 GPU with 24 GB of dedicated memory and a Core i9-13900KF CPU with 32 GB of RAM.

### Evaluation strategy

Common segmentation evaluation metrics, including Dice coefficient, Jaccard index, precision, sensitivity, specificity, accuracy, mean surface distance and segment volume difference were used to compare the predicted segmentation with the reference ones. The Mann-Whitney U test was employed to compare the models’ performance on NC and ASC images. In other words, we compared performance between tasks#1 and #2 as well as between tasks #3 and #4, seeking statistically significant differences using a two-tailed P-value of 0.05 as the threshold.

## Results

### Tasks #1 and #2

For tasks #1 and #2, an average Dice value over all organs of 0.81 ± 0.15 and 0.82 ± 0.14 was achieved, respectively. As shown in Figure 3, in terms of Dice scores, task #1 demonstrated superior performance across most organs compared to task #2.

**Figure 3.**
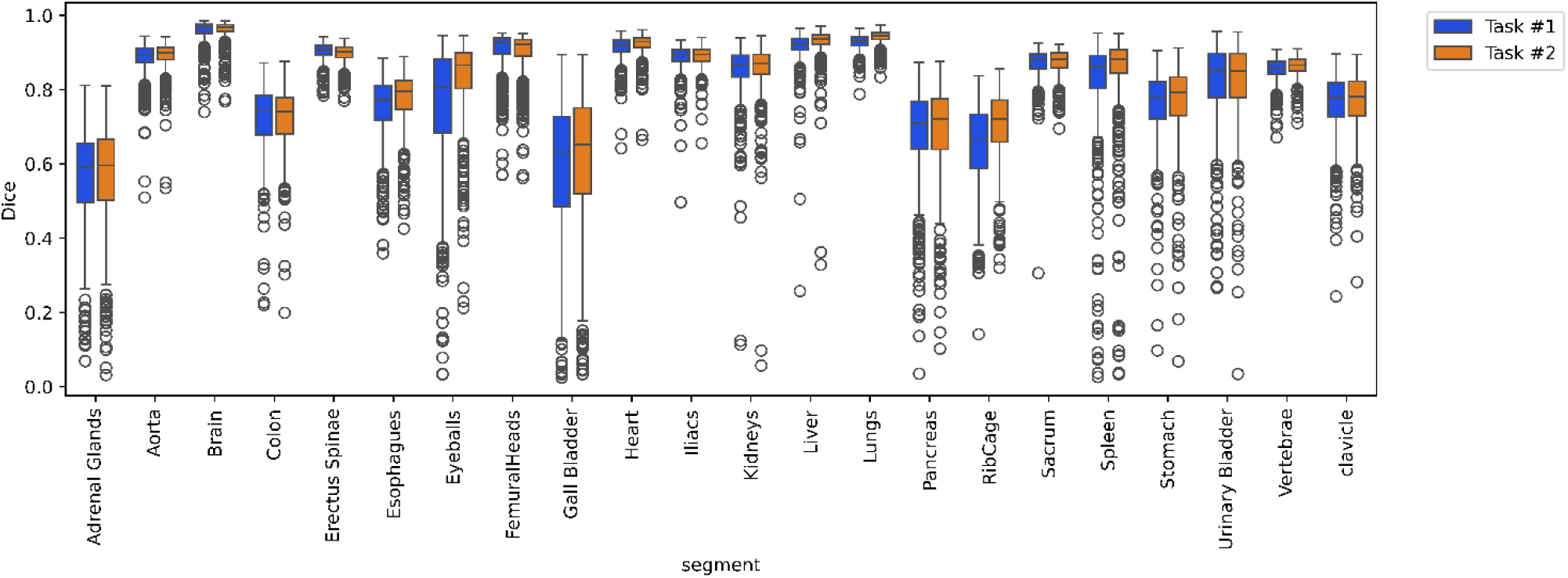
Box plot of Dice coefficients for task #1 vs #2.

Table 3 and Table 4 summarize the details of five-fold cross-validation results for tasks #1 and #2, respectively. The highest Dice coefficients were achieved for the Brain and Lungs, while the lowest values were found for smaller organs, such as AGs. The detailed results separated by every fold may be found in supplementary Table 1.

**Table 3.** Average performance metrics of our models for Task #1 from five-fold cross-validation.

| Task | Segment | Dice | Jaccard | Sensitivity | Specificity | Precision | Accuracy | Mean Surface Distance (mm) | Volume Difference (mL) |
| --- | --- | --- | --- | --- | --- | --- | --- | --- | --- |
| Task #1 | AG | 0.561 ± 0.135 | 0.402 ± 0.122 | 0.572 ± 0.157 | 1.0 ± 0.0 | 0.579 ± 0.146 | 1.0 ± 0.0 | 2.231 ± 2.006 | -0.309 ± 2.588 |
|  | Aorta | 0.884 ± 0.045 | 0.796 ± 0.066 | 0.888 ± 0.051 | 1.0 ± 0.0 | 0.883 ± 0.054 | 1.0 ± 0.0 | 1.023 ± 0.629 | -0.52 ± 26.031 |
|  | brain | 0.958 ± 0.032 | 0.92 ± 0.054 | 0.956 ± 0.032 | 1.0 ± 0.0 | 0.96 ± 0.041 | 1.0 ± 0.0 | 6.631 ± 14.308 | -7.448 ± 64.478 |
|  | clavicles | 0.762 ± 0.084 | 0.622 ± 0.101 | 0.767 ± 0.092 | 1.0 ± 0.0 | 0.761 ± 0.094 | 1.0 ± 0.0 | 2.529 ± 4.705 | -0.055 ± 9.39 |
|  | Colon | 0.725 ± 0.091 | 0.576 ± 0.103 | 0.711 ± 0.1 | 1.0 ± 0.0 | 0.745 ± 0.095 | 0.999 ± 0.0 | 4.062 ± 2.404 | -48.169 ± 141.816 |
|  | Esophagus | 0.754 ± 0.085 | 0.612 ± 0.101 | 0.763 ± 0.096 | 1.0 ± 0.0 | 0.751 ± 0.094 | 1.0 ± 0.0 | 1.295 ± 0.857 | 0.528 ± 7.615 |
|  | Eyeballs | 0.757 ± 0.171 | 0.635 ± 0.192 | 0.779 ± 0.171 | 1.0 ± 0.0 | 0.742 ± 0.177 | 1.0 ± 0.0 | 1.879 ± 2.475 | 1.14 ± 2.745 |
|  | Femoral Heads | 0.905 ± 0.058 | 0.831 ± 0.086 | 0.914 ± 0.047 | 1.0 ± 0.0 | 0.9 ± 0.079 | 1.0 ± 0.0 | 3.607 ± 9.742 | 22.545 ± 99.621 |
|  | GB | 0.586 ± 0.193 | 0.439 ± 0.182 | 0.596 ± 0.21 | 1.0 ± 0.0 | 0.625 ± 0.214 | 1.0 ± 0.0 | 5.102 ± 8.522 | -2.832 ± 14.219 |
|  | Heart | 0.913 ± 0.034 | 0.841 ± 0.053 | 0.918 ± 0.049 | 1.0 ± 0.0 | 0.911 ± 0.048 | 1.0 ± 0.0 | 3.038 ± 5.685 | 3.421 ± 64.693 |
|  | Hips | 0.887 ± 0.036 | 0.799 ± 0.053 | 0.886 ± 0.039 | 1.0 ± 0.0 | 0.889 ± 0.045 | 1.0 ± 0.0 | 0.809 ± 0.386 | -4.637 ± 34.58 |
|  | Kidneys | 0.851 ± 0.077 | 0.747 ± 0.095 | 0.866 ± 0.084 | 1.0 ± 0.0 | 0.844 ± 0.087 | 1.0 ± 0.0 | 2.164 ± 3.047 | 5.254 ± 49.649 |
|  | Liver | 0.915 ± 0.049 | 0.846 ± 0.067 | 0.92 ± 0.063 | 1.0 ± 0.0 | 0.914 ± 0.052 | 1.0 ± 0.0 | 2.48 ± 1.942 | 3.991 ± 222.317 |
|  | Lungs | 0.927 ± 0.022 | 0.865 ± 0.037 | 0.929 ± 0.034 | 1.0 ± 0.0 | 0.927 ± 0.038 | 0.999 ± 0.0 | 1.824 ± 0.841 | 4.763 ± 214.312 |
|  | Pancreas | 0.682 ± 0.126 | 0.53 ± 0.131 | 0.688 ± 0.143 | 1.0 ± 0.0 | 0.691 ± 0.131 | 1.0 ± 0.0 | 3.561 ± 3.175 | -1.579 ± 19.486 |
|  | RAM | 0.903 ± 0.025 | 0.824 ± 0.041 | 0.902 ± 0.03 | 1.0 ± 0.0 | 0.906 ± 0.036 | 1.0 ± 0.0 | 1.083 ± 0.387 | -9.367 ± 54.561 |
|  | Rib Cage | 0.651 ± 0.111 | 0.492 ± 0.116 | 0.662 ± 0.115 | 1.0 ± 0.0 | 0.645 ± 0.12 | 0.999 ± 0.0 | 1.729 ± 1.758 | 10.792 ± 68.369 |
|  | Sacrum | 0.872 ± 0.042 | 0.776 ± 0.056 | 0.888 ± 0.046 | 1.0 ± 0.0 | 0.859 ± 0.05 | 1.0 ± 0.0 | 1.237 ± 0.791 | 8.624 ± 16.751 |
|  | Spleen | 0.819 ± 0.141 | 0.711 ± 0.153 | 0.834 ± 0.148 | 1.0 ± 0.0 | 0.82 ± 0.142 | 1.0 ± 0.0 | 3.581 ± 6.563 | 4.592 ± 58.892 |
|  | Stomach | 0.758 ± 0.1 | 0.62 ± 0.113 | 0.763 ± 0.118 | 1.0 ± 0.0 | 0.767 ± 0.106 | 1.0 ± 0.0 | 3.637 ± 3.425 | -3.837 ± 43.979 |
|  | UB | 0.821 ± 0.112 | 0.71 ± 0.141 | 0.832 ± 0.137 | 1.0 ± 0.0 | 0.835 ± 0.138 | 1.0 ± 0.0 | 3.774 ± 15.654 | -4.205 ± 44.513 |
|  | Vertebrae | 0.856 ± 0.031 | 0.75 ± 0.046 | 0.862 ± 0.035 | 1.0 ± 0.0 | 0.852 ± 0.04 | 1.0 ± 0.0 | 1.306 ± 2.694 | 11.017 ± 53.781 |

**Table 4.**
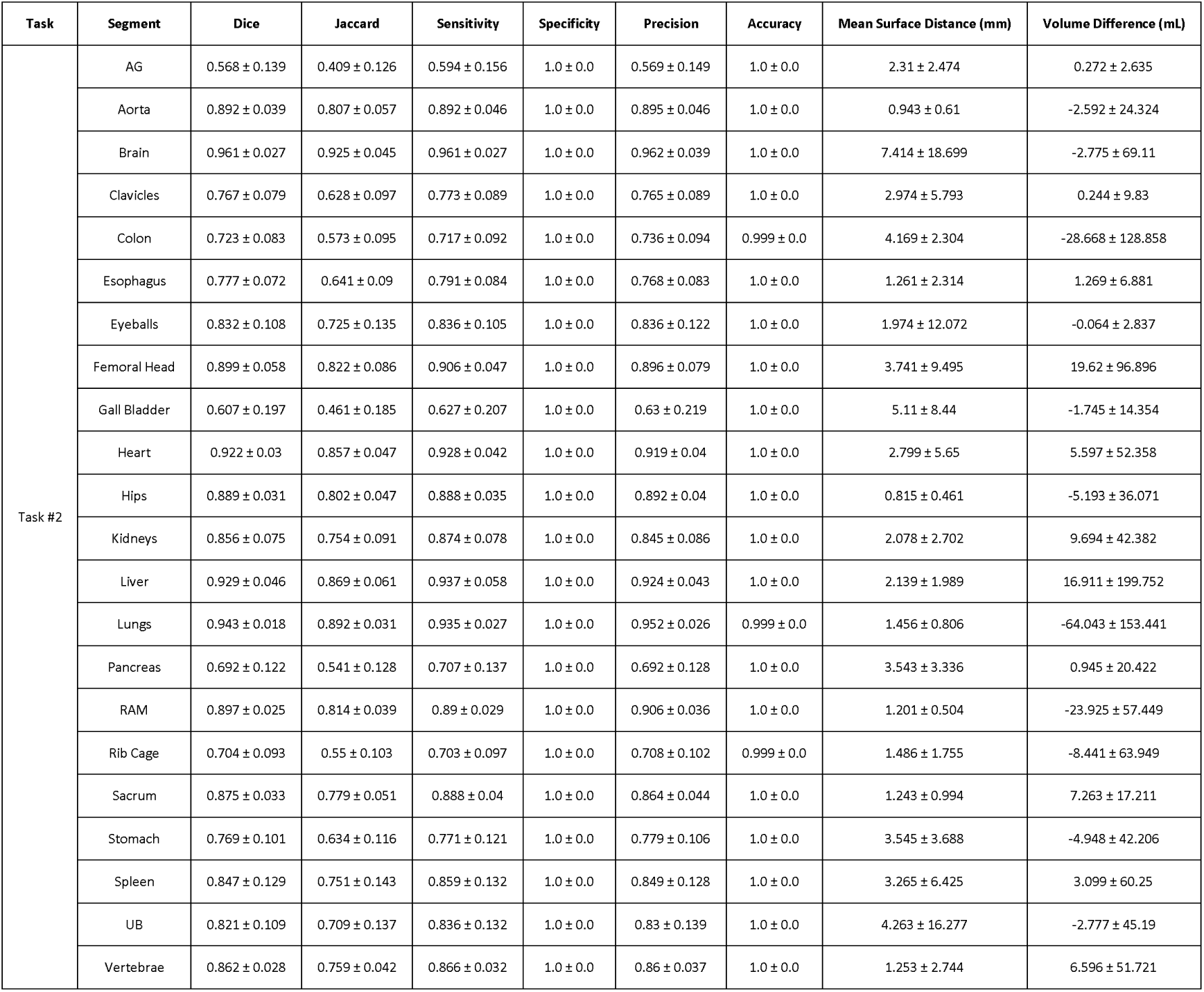
Evaluation metrics for Task #2, averaged over five-fold cross-validation.

Table ***5*** compares P-Values between tasks #1 and #2, indicating significant differences across most organs. There are significant differences in Dice values for most organs between tasks #1 and #2, while volume differences show significance in fewer organs. An example of our model output for task #2 tested on a noisy dynamic acquisition is shown in supplementary figure 2.

**Table 5.** Mann-Whitney P-values comparing task #1 and task #2. P<0.05 reflects statistically significant difference.

| Organ | Dice | Jaccard | Mean Surface Distance | Volume Difference |
| --- | --- | --- | --- | --- |
| Liver | 0.000 | 0.000 | 0.000 | 0.089 |
| brain | 0.619 | 0.619 | 0.485 | 0.164 |
| AG | 0.236 | 0.236 | 0.315 | 0.001 |
| Stomach | 0.011 | 0.011 | 0.069 | 0.473 |
| Rib Cage | 0.000 | 0.000 | 0.000 | 0.000 |
| Colon | 0.396 | 0.396 | 0.136 | 0.012 |
| Erectus Spinae | 0.000 | 0.000 | 0.000 | 0.000 |
| Sacrum | 0.318 | 0.318 | 0.248 | 0.255 |
| Aorta | 0.003 | 0.003 | 0.003 | 0.024 |
| Clavicle | 0.385 | 0.385 | 0.887 | 0.633 |
| Esophagus | 0.000 | 0.000 | 0.000 | 0.068 |
| Vertebrae | 0.001 | 0.001 | 0.006 | 0.092 |
| Eyeballs | 0.000 | 0.000 | 0.000 | 0.000 |
| Femoral Head | 0.000 | 0.000 | 0.004 | 0.342 |
| GB | 0.034 | 0.034 | 0.261 | 0.061 |
| Spleen | 0.000 | 0.000 | 0.000 | 0.458 |
| Kidneys | 0.162 | 0.162 | 0.296 | 0.263 |
| Lungs | 0.000 | 0.000 | 0.000 | 0.000 |
| Hips | 0.421 | 0.421 | 0.763 | 0.855 |
| Pancreas | 0.175 | 0.175 | 0.350 | 0.022 |
| UB | 0.741 | 0.741 | 0.362 | 0.463 |
| Heart | 0.000 | 0.000 | 0.000 | 0.907 |

Figure 4 demonstrates an example of segmented organs for tasks #1 and #2 on a case with a good match between PET and CT images from a cross-validation strategy, depicting the excellent performance of our models.

**Figure 4.**
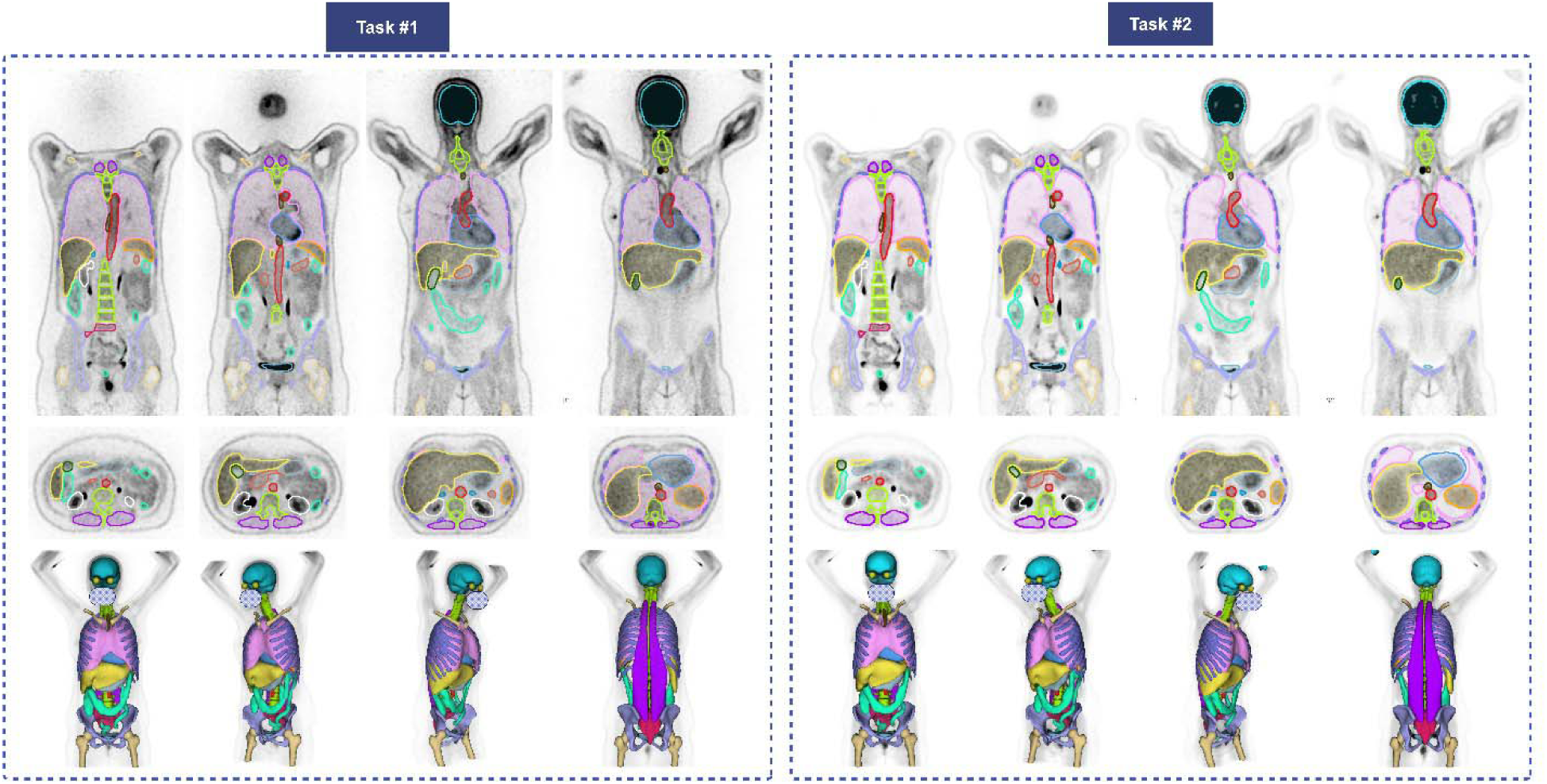
Coronal (top), axial (middle), and 3D (bottom) visualizations of the segmentations for tasks #1 and #2. Each color presents one organ; the internal organs, such as kidneys, are not visible in 3D-rendered images. The face is masked for privacy.

Figure 5 shows an example of PET/CT image with unreliable CT segmentation due to respiratory mismatch affecting the segmentation of moving organs, especially the Lungs, Liver, and Spleen. This case was excluded from cross-validation training, the trained models of tasks#1 and #2 were ensembled on the corresponding images. In other words, task#1 model was tested on PET-NC and task#2 model on PET-ASC images.

**Figure 5.**
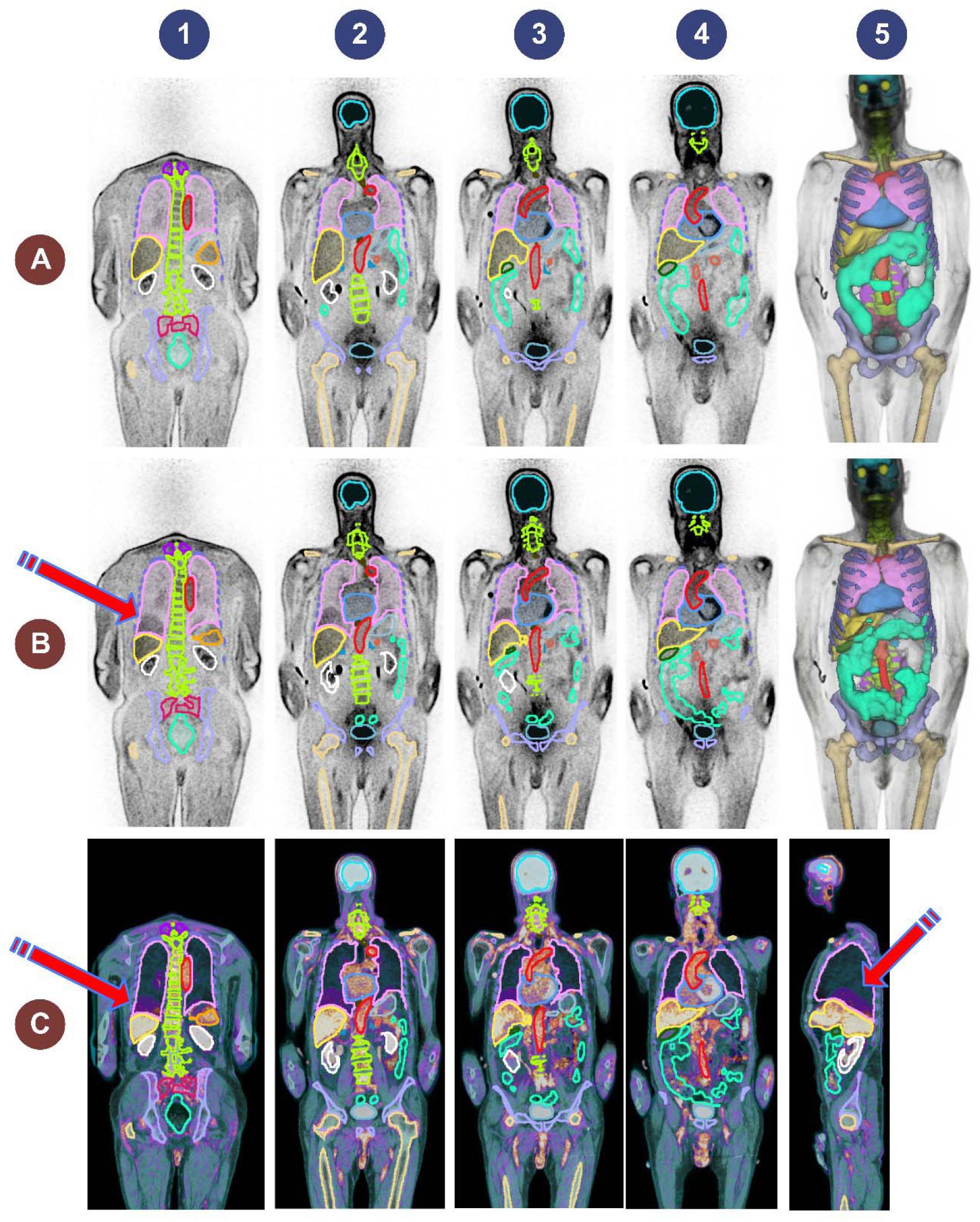
Representative excluded case presenting an image with respiratory mismatch between PET and CT images. A (top row): PET-NC image and task #1 generated masks, columns 1 to 4 show coronal images whereas column 5 shows the 3D rendered segmentations. B (middle row): PET-NC image and the segmentation masks generated on the co-registered CTAC image, columns 1 to 4 show coronal images whereas column 5 shows the 3D rendered segmentations. The arrow shows the mismatch at lung/liver interface. C (bottom row): the fused PET-NC and CT in coronal (columns 1 to 4) and sagittal (column 5) views. The arrows highlight the mismatch regions.

### Tasks #3 and #4

The average of the Dice scores over 15 organs from five-fold cross-validation were 0.766 ± 0.171 and 0.788 ± 0.163 for tasks #3 and #4, respectively. The performance metrics for tasks #3 and #4 are summarized in Table 6 and Table 7. The detailed performance metrics are reported separately for each fold in supplementary Table 1. The Dice values for task #4 were significantly higher than those for task #3 with P-Values below 0.05 for most organs, except for larger organs with a clear objective contrast on ^68^Ga-NC images, such as the Hips, Sacrum, Vertebrae, Kidneys, and UB. The P-Values are reported in Table 8. The lowest Dice value was observed for AG whereas the highest was achieved for the Brain. Figure 6 displays the box plot of Dice scores for the included organs in tasks #3 and #4.

**Table 6.**
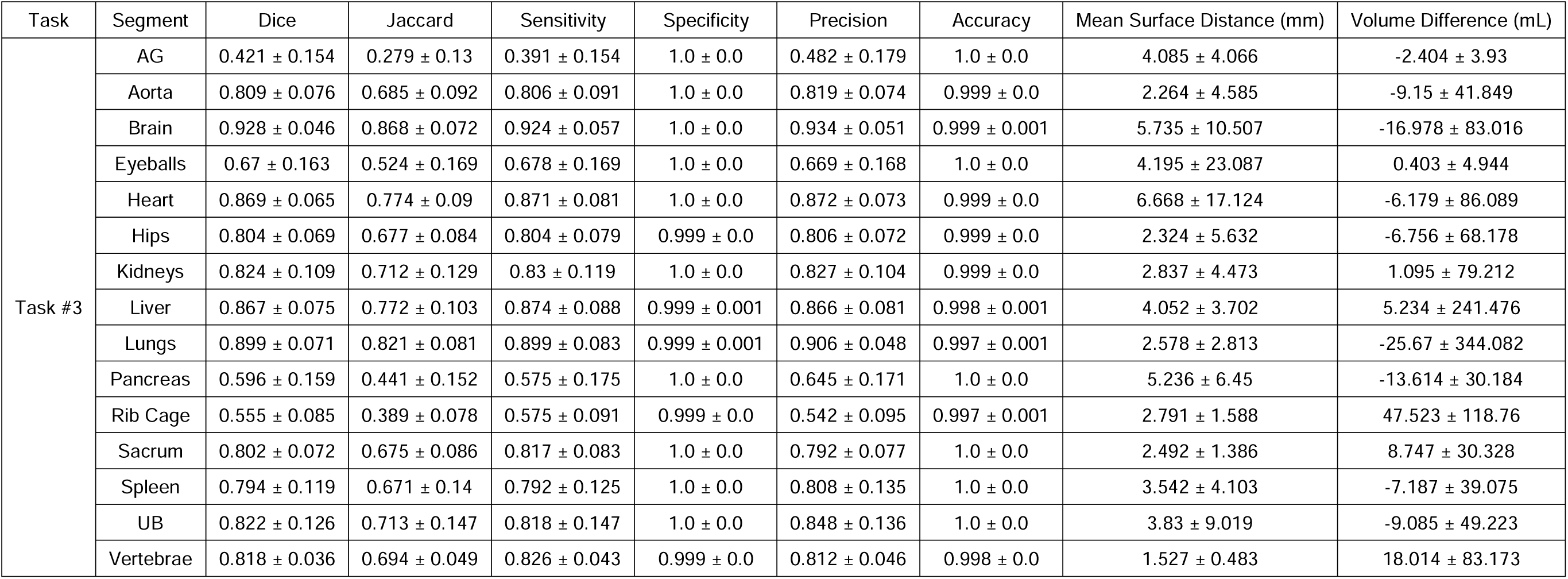
Average segmentation metrics from five-fold cross-validation for Task #3 for all included organs.

**Table 7.** Performance metrics from five-fold cross-validation for Task #4.

| Task | Segment | Dice | Jaccard | Sensitivity | Specificity | Precision | Accuracy | Mean Surface Distance (mm) | Volume Difference (mL) |
| --- | --- | --- | --- | --- | --- | --- | --- | --- | --- |
| Task #4 | AG | 0.437 ± 0.155 | 0.293 ± 0.133 | 0.414 ± 0.154 | 1.0 ± 0.0 | 0.485 ± 0.179 | 1.0 ± 0.0 | 3.355 ± 2.967 | -1.98 ± 3.422 |
|  | Aorta | 0.825 ± 0.075 | 0.708 ± 0.096 | 0.824 ± 0.081 | 1.0 ± 0.0 | 0.832 ± 0.085 | 1.0 ± 0.0 | 1.913 ± 2.273 | -3.939 ± 69.773 |
|  | Brain | 0.942 ± 0.042 | 0.893 ± 0.064 | 0.942 ± 0.053 | 1.0 ± 0.0 | 0.945 ± 0.044 | 0.999 ± 0.0 | 5.559 ± 11.292 | -9.794 ± 84.242 |
|  | Eyeballs | 0.753 ± 0.134 | 0.62 ± 0.151 | 0.747 ± 0.132 | 1.0 ± 0.0 | 0.774 ± 0.14 | 1.0 ± 0.0 | 6.797 ± 37.747 | 0.379 ± 15.853 |
|  | Heart | 0.897 ± 0.057 | 0.818 ± 0.081 | 0.898 ± 0.073 | 1.0 ± 0.0 | 0.9 ± 0.061 | 0.999 ± 0.0 | 6.186 ± 17.138 | -5.302 ± 73.578 |
|  | Hips | 0.812 ± 0.068 | 0.688 ± 0.082 | 0.803 ± 0.08 | 0.999 ± 0.0 | 0.824 ± 0.067 | 0.999 ± 0.0 | 2.189 ± 6.148 | -25.21 ± 70.011 |
|  | Kidneys | 0.819 ± 0.12 | 0.707 ± 0.136 | 0.824 ± 0.118 | 1.0 ± 0.0 | 0.826 ± 0.126 | 0.999 ± 0.001 | 3.641 ± 8.759 | 5.333 ± 132.237 |
|  | Liver | 0.904 ± 0.062 | 0.829 ± 0.087 | 0.906 ± 0.082 | 0.999 ± 0.0 | 0.906 ± 0.055 | 0.999 ± 0.001 | 2.599 ± 2.166 | -9.704 ± 222.004 |
|  | Lungs | 0.926 ± 0.035 | 0.864 ± 0.054 | 0.926 ± 0.047 | 0.999 ± 0.001 | 0.927 ± 0.037 | 0.998 ± 0.001 | 1.744 ± 1.395 | -18.3 ± 226.443 |
|  | Pancreas | 0.63 ± 0.151 | 0.476 ± 0.148 | 0.617 ± 0.164 | 1.0 ± 0.0 | 0.665 ± 0.17 | 1.0 ± 0.0 | 4.417 ± 5.866 | -9.752 ± 28.548 |
|  | Rib Cage | 0.585 ± 0.077 | 0.417 ± 0.072 | 0.607 ± 0.081 | 0.999 ± 0.0 | 0.571 ± 0.092 | 0.997 ± 0.001 | 2.391 ± 1.414 | 49.412 ± 122.563 |
|  | Sacrum | 0.793 ± 0.086 | 0.664 ± 0.101 | 0.804 ± 0.095 | 1.0 ± 0.0 | 0.786 ± 0.091 | 1.0 ± 0.0 | 3.595 ± 10.435 | 6.662 ± 33.149 |
|  | Spleen | 0.839 ± 0.097 | 0.732 ± 0.123 | 0.838 ± 0.107 | 1.0 ± 0.0 | 0.847 ± 0.106 | 1.0 ± 0.0 | 2.752 ± 5.183 | -2.885 ± 31.768 |
|  | UB | 0.834 ± 0.11 | 0.728 ± 0.135 | 0.828 ± 0.138 | 1.0 ± 0.0 | 0.861 ± 0.125 | 1.0 ± 0.0 | 2.628 ± 2.176 | -9.362 ± 46.993 |
|  | Vertebrae | 0.819 ± 0.039 | 0.696 ± 0.053 | 0.831 ± 0.05 | 0.999 ± 0.0 | 0.81 ± 0.048 | 0.998 ± 0.001 | 1.54 ± 0.598 | 29.778 ± 96.0 |

**Table 8.** P-values of Mann-Whitney statistical test comparing the performance metrics for task#3 vs task #4.

| Organ | P-Value |  |  |  |
| --- | --- | --- | --- | --- |
|  | Dice | Jaccard | Mean Surface Distance | Volume Difference |
| AG | 0.269 | 0.269 | 0.049 | 0.210 |
| Aorta | 0.000 | 0.000 | 0.001 | 0.962 |
| Brain | 0.000 | 0.000 | 0.015 | 0.083 |
| Eyeballs | 0.000 | 0.000 | 0.000 | 0.000 |
| Hips | 0.141 | 0.141 | 0.054 | 0.004 |
| Kidneys | 0.668 | 0.668 | 0.776 | 0.373 |
| Liver | 0.000 | 0.000 | 0.000 | 0.359 |
| Lungs | 0.000 | 0.000 | 0.000 | 0.684 |
| Pancreas | 0.012 | 0.012 | 0.004 | 0.331 |
| Rib Cage | 0.000 | 0.000 | 0.000 | 0.909 |
| Sacrum | 0.565 | 0.565 | 0.532 | 0.375 |
| Spleen | 0.000 | 0.000 | 0.000 | 0.607 |
| UB | 0.359 | 0.359 | 0.135 | 0.749 |
| Vertebrae | 0.684 | 0.684 | 0.837 | 0.164 |
| Heart | 0.000 | 0.000 | 0.000 | 0.979 |

**Figure 6.**
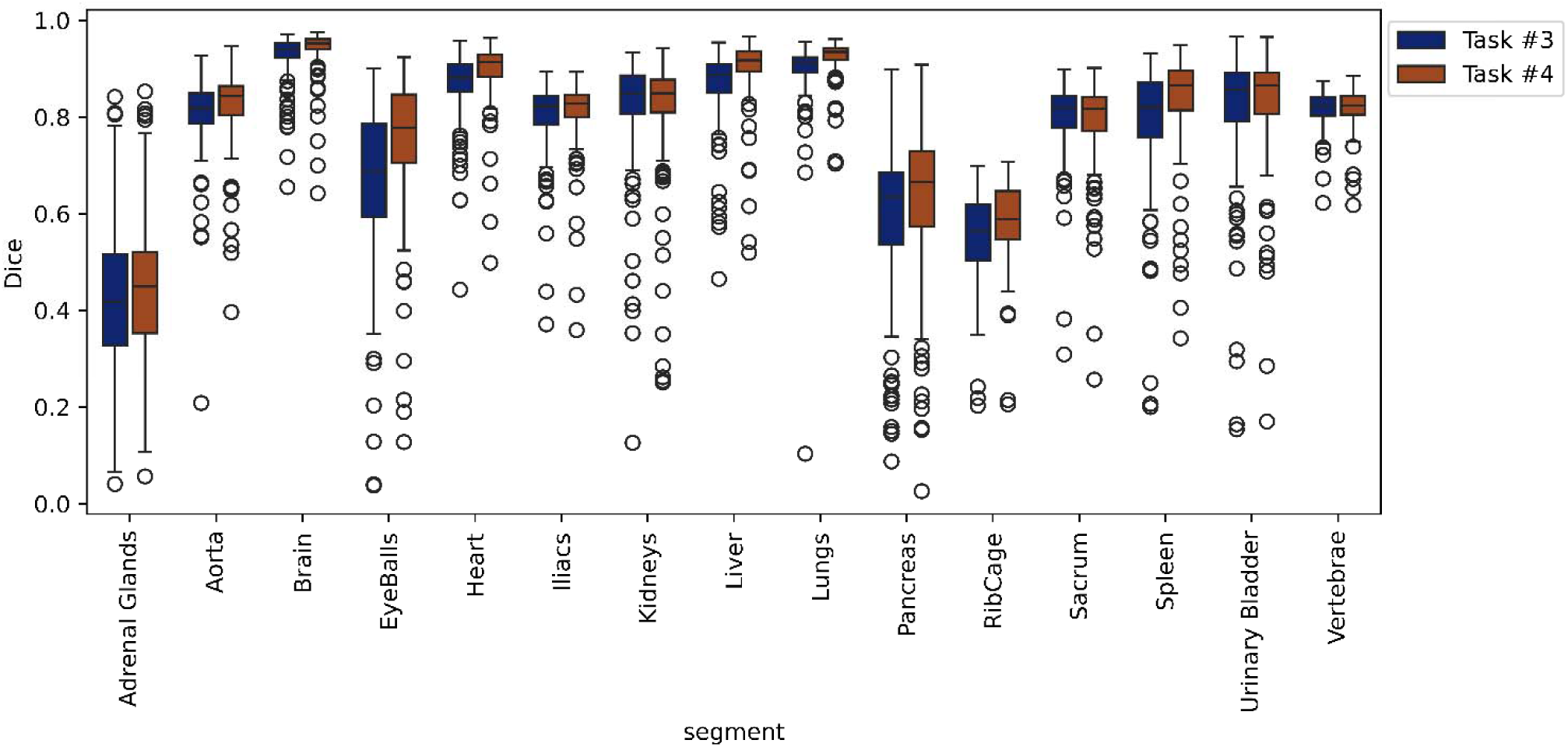
Box plots showing the Dice scores for task #3 and #4 for every 15 included organs.

Figure 7 illustrates an example with strong alignment between CT and PET within the 5-fold cross-validation data split for tasks #3 and #4.

**Figure 7.**
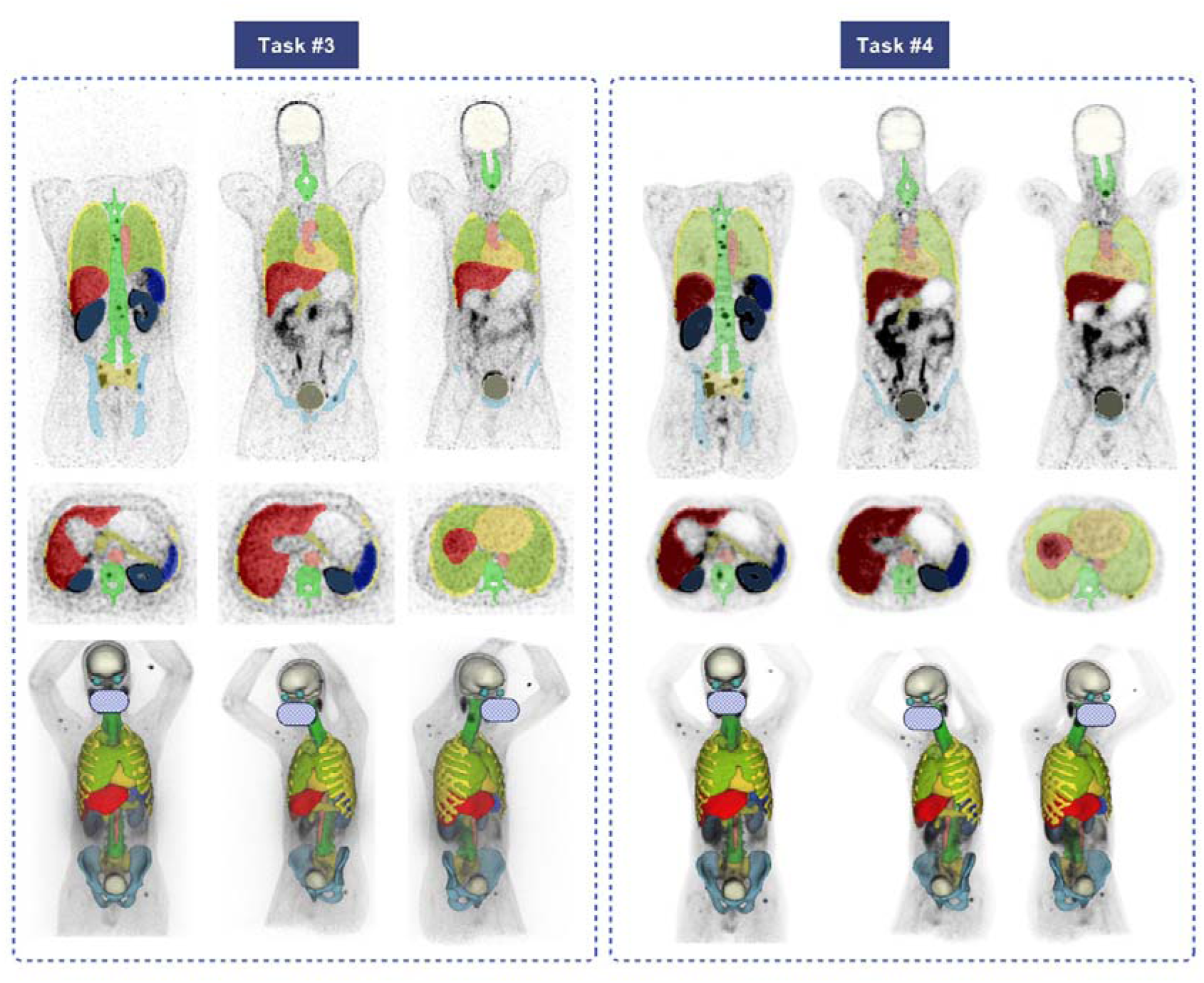
Representative case with segmentations generated on PET-NC (task #3) and PET-ASC images (task #4). Top row: coronal slices, middle row: axial and bottom row: 3D rendered segmentations. Face is masked for privacy.

Figure 8 depicts an image from the excluded studies demonstrating the mismatch between PET and CT images. This case shows the unreliable CT generated masks and the excellent performance of our model in delineating organs.

**Figure 8.**
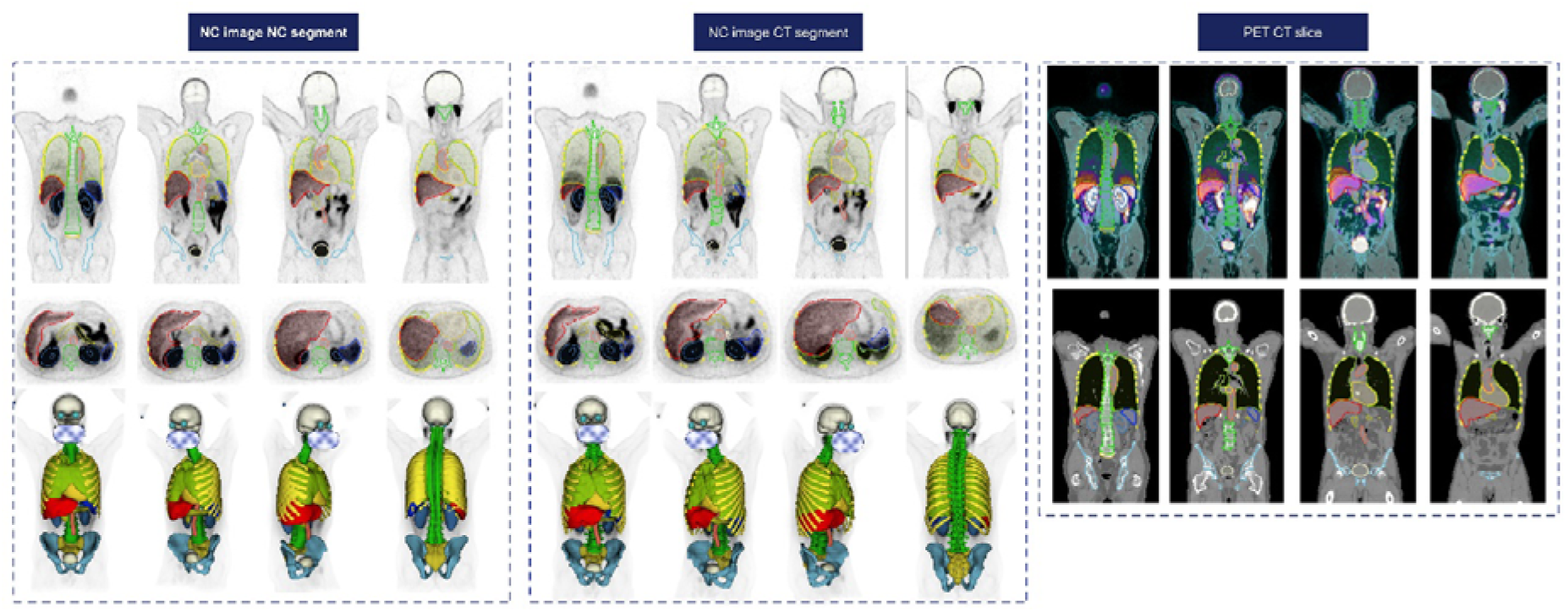
^68^Ga-PSMA PET/CT image with respiratory mismatch. It should be noted that the kidney boundaries are expanded in the visualized images due tNo selected window width/level. Face is masked for privacy.

## Discussion

Automatic, fast, and accurate segmentation of medical images has become one of the hottest topics in precision medicine for personalized dosimetry and image quantification [41–45]. In nuclear medicine, where PET/CT and SPECT/CT are commonly performed for diagnostic and therapeutic purposes, automated organ segmentation is crucial. Recent studies showed the importance of Organomics and organ information in overall survival prediction [15; 20]. as well as pre-therapy dose prediction in PSMA theragnostic procedures [14]. The common method used for automated organ delineation in hybrid PET/CT and SPECT/CT relies on the co-registered CT images. However, this approach is subject to limitations, such as the highly prevalent mismatch between emission and CT images and the low quality of low-dose CTAC images. Urinary bladder filling and bowel movements are inevitable and could cause more problematic mismatch between the CT generated masks and realistic organ position, shape and size depicted on PET images. Additionally, not all PET scanners are equipped with CT for attenuation and scatter correction, and as such, an approach that doesn’t rely on CT is necessary.

We targeted two commonly used tracers for PET imaging and developed a comprehensive DL segmentation pipeline for the automated delineation of multiple organs to be used in different clinical scenarios. To ensure a strong match between emission and CT images in the training set and to prevent the trained DL models from being affected by mismatch, we first cleaned our data by excluding PET/CT image pairs presenting with respiratory, cardiac, and bulk motion mismatches. After visual assessment of our initially included dataset, we excluded more than 65% of the 2062 images from our study, emphasizing the high prevalence of mismatch in PET/CT imaging.

We included different numbers of organs depending on the tracer as ^68^Ga-PSMA PET images contain less anatomical information. Our goal was to develop a reliable model based on clinical studies for potential implementation in clinical setting. The first scenario for using our models involve performing segmentation on PET ASC images corrected either by CT or any other method, such as MLAA or DL-based ASC techniques. In this scenario, the CT image is either unavailable or too noisy to be segmented through DL . To address this limitation, we provided models from tasks #2 and #4 for delineation on PET ASC images, as they outperformed the models of tasks #1 and #3, which use NC images. PET ASC images benefit from better contrast and contain more information due to corrections for degrading factors, such as attenuation, scatter and point-spread function (PSF).

Additionally, we considered the second scenario of performing CT-less PET segmentation where the PET ASC image is corrected with a mismatched CT, or PET-ASC images are not available. Such corrections can induce unacceptable mismatch artifacts on PET ASC images, removing the useful information e.g., in areas such as chest abdomen interval or causing halo artifacts which are very common in ^68^Ga-PSMA PET/CT imaging. In this scenario, as shown in Figure 5 and Figure 8, the chest area was affected, and the DL model trained on PET ASC images only on cases without mismatch, may identify it as lung tissue. To address this issue, we implemented two strategies including tasks #1 and #3 to provide a reliable segmentation solution for all potential clinical scenarios. The performance of our models in the second scenario is lower than those in the first scenario as the NC images suffer from multiple artifacts, and are not corrected for attenuation and scatter, and usually do not include time-of-flight (TOF) and PSF correction. We hypothesize that this would be a versatile solution by considering real clinical needs and could tackle the issue more effectively.

We employed state-of-the-art nnU-Net V2 pipeline, which has shown promising results in recent medical image segmentation studies. Our model achieved excellent accuracy in segmenting organs, such as the Lungs, Brain, and Liver. However, it achieved lower performance in a few smaller organs with lower objective contrast and visibility in PET images, especially when using NC images as input, such as AGs. The overall performance was superior in tasks #1 and #2 using FDG PET images compared to tasks #3 and #4 using ^68^Ga-PET, as anticipated. The difference can be attributed to the lower structural information in ^68^Ga-PET images because of specific uptake patterns of ^68^Ga-PSMA tracer. As shown in supplementary figure 2, our models have shown acceptable performance even on very noisy dynamic FDG PET images. It should be noted that these images are acquired shortly after tracer injection, thus a different radiopharmaceutical uptake pattern can be observed compared to delayed PET images (usually acquired around 60 minutes post-injection). Despite this fact, our model showed robust performance on dynamic, noisy frames.

While the Dice coefficient alone may have limitations for evaluation of image segmentation performance [46], we extensively evaluated the performance of our models using multiple metrics, including Dice, Jaccard, mean surface distance, and the volume difference between the reference and the predicted masks. Our study achieved significantly better results compared to the study by Yazdani et al. [47] where few organs were included for segmentation. Our Dice scores were 0.82 vs. 0.80, 0.90 vs. 0.88, 0.84 vs. 0.79, and 0.83 vs. 0.81 for kidneys, liver, spleen, and urinary bladder organs in task #4. The improved performance could be due to our approach of excluding cases with mismatches, which could mislead the DL model during training and underestimate the Dice value when unreliable segmentation masks are used as reference in those cases. Klyuzhin et al. [48] developed a multi-organ segmentation model for ^68^Ga-PSMA images using both PET and CT images as inputs in their UNET model. Our model, however, utilizes only the emission images to overcome the aforementioned limitations.

This work inherently bears a number of limitations. First, we developed PET organ segmentation models for two common tracers and suggested training new models for other tracers. However, transfer learning is one option that should be considered. CT segmentation does not share this dependency on the tracer used. Another limitation of our study is the limited number of training dataset acquired on only two PET/CT scanners from the same manufacturer., Potential end-users would need to perform finetuning on our publicly available models using their own local datasets.

## Conclusion

We developed DL-powered CT-less automated organ segmentation models from PET images for two common tracers used in PET/CT imaging to overcome the limitations of CT segmentation in delineation and quantification. Our model showed acceptable performance; however, the models using PET-ASC images as input achieved a better performance. The method can be used in the clinical to enable a number of clinical and research applications.

## Code and data availability

These models and inference instructions will be available on GitHub upon publication of this article.

## Ethical approval

The study was approved by the Ethics committee of the Canton of Geneva, Switzerland. Consent forms were waived owing to the retrospective nature of the study.

## Data Availability

Datasets are not publicly available.
The nnU-Net trained models will be available on Github oage of the first author.

## Abbreviations

PET: Positron Emission Tomography
CT: Computed Tomography
NC: Non-Corrected
ASC: Attenuation and Scatter Corrected
PSMA: Prostate Specific Membrane
Antigen FDG: Fluorodeoxyglucose
NET: Neuroendocrine tumor
UB: Urinary Bladder
AG: Adrenal Glands
CTDI_vol_: Volumetric Computed Tomography Dose Index

## References

1 Zaidi H, El Naqa I (2010) PET-guided delineation of radiation therapy treatment volumes: a survey of image segmentation techniques. European Journal of Nuclear Medicine and Molecular Imaging 37:2165–2187

2 Ferrari M, Travaini LL, Ciardo D et al (2017) Interim (18)FDG PET/CT during radiochemotherapy in the management of pelvic malignancies: A systematic review. Crit Rev Oncol Hematol 113:28–42

3 Spence AM, Muzi M, Mankoff DA, et al (2004) 18F-FDG PET of gliomas at delayed intervals: improved distinction between tumor and normal gray matter. J Nucl Med 45:1653–1659

4 Qin C, Shao F, Hu F et al (2020) 18F-FDG PET/CT in diagnostic and prognostic evaluation of patients with cardiac masses: a retrospective study. European journal of nuclear medicine and molecular imaging 47:1083–1093

5 Rai BP, Baum RP, Patel A et al (2016) The Role of Positron Emission Tomography With (68)Gallium (Ga)-Labeled Prostate-specific Membrane Antigen (PSMA) in the Management of Patients With Organ-confined and Locally Advanced Prostate Cancer Prior to Radical Treatment and After Radical Prostatectomy. Urology 95:11–15

6 Hu X, Wu Y, Yang P, Wang J, Wang P, Cai J (2022) Performance of 68Ga-labeled prostate-specific membrane antigen ligand positron emission tomography/computed tomography in the diagnosis of primary prostate cancer: a systematic review and meta-analysis. Int Braz J Urol 48:891–902

7 Buteau JP, Martin AJ, Emmett L et al (2022) PSMA and FDG-PET as predictive and prognostic biomarkers in patients given [(177)Lu]Lu-PSMA-617 versus cabazitaxel for metastatic castration-resistant prostate cancer (TheraP): a biomarker analysis from a randomised, open-label, phase 2 trial. Lancet Oncol 23:1389–1397

8 Zhuang M, García DV, Kramer GM et al (2019) Variability and repeatability of quantitative uptake metrics in 18F-FDG PET/CT of non–small cell lung cancer: Impact of segmentation method, uptake interval, and reconstruction protocol. Journal of Nuclear Medicine 60:600–607

9 Takahashi ME, Mosci C, Souza EM et al (2019) Proposal for a quantitative 18F-FDG PET/CT metabolic parameter to assess the intensity of bone involvement in multiple myeloma. Scientific reports 9:16429

10 Ziai P, Hayeri MR, Salei A et al (2016) Role of optimal quantification of FDG PET imaging in the clinical practice of radiology. Radiographics 36:481–496

11 Zaidi H, Karakatsanis N (2018) Towards enhanced PET quantification in clinical oncology. Br J Radiol 91:20170508

12 Pandit-Taskar N, Iravani A, Lee D et al (2021) Dosimetry in clinical radiopharmaceutical therapy of cancer: practicality versus perfection in current practice. Journal of Nuclear Medicine 62:60S–72S

13 Dewaraja YK, Covert EC, Fitzpatrick K, et al (2022) Intra-and Inter-operator variability in manual tumor segmentation: Impact on radionuclide therapy dosimetry.

14 Xue S, Gafita A, Zhao Y et al (2024) Pre-therapy PET-based voxel-wise dosimetry prediction by characterizing intra-organ heterogeneity in PSMA-directed radiopharmaceutical theranostics. European Journal of Nuclear Medicine and Molecular Imaging. 10.1007/s00259-024-06737-3

15 Kibrom BG, Anne-Ségolène C, Laetitia V et al (2023) Tumor Location Relative to the Spleen Is a Prognostic Factor in Lymphoma Patients: A Demonstration from the REMARC Trial. Journal of Nuclear Medicine. 10.2967/jnumed.123.266322:jnumed.123.266322

16 Sharma R, Wang WM, Yusuf S et al (2019) (68)Ga-DOTATATE PET/CT parameters predict response to peptide receptor radionuclide therapy in neuroendocrine tumours. Radiother Oncol 141:108–115

17 Ortega C, Wong RKS, Schaefferkoetter J et al (2021) Quantitative (68)Ga-DOTATATE PET/CT Parameters for the Prediction of Therapy Response in Patients with Progressive Metastatic Neuroendocrine Tumors Treated with (177)Lu-DOTATATE. J Nucl Med 62:1406–1414

18 Mansouri Z SY, Hajianfar G, Bianchetto Wolf N, Knappe L, Xhepa G, Gleyzolle A, Ricoeur A, Garibott V, Mainta I, Zaidi H. (2024) The role of biomarkers and dosimetry parameters in overall and progression free survival prediction for patients treated with personalized 90Y glass microspheres SIRT: a preliminary machine learning study. Eur J Nucl Med Mol Imaging 51:in press.

19 Mansouri Z, Salimi Y, Amini M et al (2024) Development and validation of survival prognostic models for head and neck cancer patients using machine learning and dosiomics and CT radiomics features: a multicentric study. Radiat Oncol 19:12

20 Salimi Y, Hajianfar G, Mansouri Z et al (2024) Organomics: A novel concept reflecting the importance of PET/CT healthy organ radiomics in non-small cell lung cancer prognosis prediction using machine learning. Clin Nucl Med 51:*in press*

21 Adam JA, Loft A, Chargari C et al (2021) EANM/SNMMI practice guideline for [(18)F]FDG PET/CT external beam radiotherapy treatment planning in uterine cervical cancer v1.0. Eur J Nucl Med Mol Imaging 48:1188–1199

22 Vaz SC, Adam JA, Delgado Bolton RC et al (2022) Joint EANM/SNMMI/ESTRO practice recommendations for the use of 2-[(18)F]FDG PET/CT external beam radiation treatment planning in lung cancer V1.0. Eur J Nucl Med Mol Imaging 49:1386–1406

23 Cornford P, van den Bergh RCN, Briers E, et al (2024) EAU-EANM-ESTRO-ESUR-ISUP-SIOG Guidelines on Prostate Cancer—2024 Update. Part I: Screening, Diagnosis, and Local Treatment with Curative Intent. European Urology. 10.1016/j.eururo.2024.03.027

24 McErlean A, Panicek DM, Zabor EC et al (2013) Intra-and interobserver variability in CT measurements in oncology. Radiology 269:451–459

25 Breen SL, Publicover J, De Silva S, et al (2007) Intraobserver and interobserver variability in GTV delineation on FDG-PET-CT images of head and neck cancers. International Journal of Radiation Oncology* Biology* Physics 68:763–770

26 Zaidi H, El Naqa I (2021) Quantitative Molecular Positron Emission Tomography Imaging Using Advanced Deep Learning Techniques. Annu Rev Biomed Eng 23:249–276

27 Wasserthal J, Breit HC, Meyer MT, et al (2023) TotalSegmentator: Robust Segmentation of 104 Anatomic Structures in CT Images. Radiol Artif Intell 5:e230024

28 Salimi Y, Shiri I, Mansouri Z, Zaidi H (2023) Deep learning-assisted multiple organ segmentation from whole-body CT images. medRxiv. 10.1101/2023.10.20.23297331:2023.2010.2020.23297331

29 Shiri I, Vafaei Sadr A, Akhavan A et al (2023) Decentralized collaborative multi-institutional PET attenuation and scatter correction using federated deep learning. Eur J Nucl Med Mol Imaging 50:1034–1050

30 Shiri I, Salimi Y, Hervier E et al (2023) Artificial Intelligence-Driven Single-Shot PET Image Artifact Detection and Disentanglement: Toward Routine Clinical Image Quality Assurance. Clin Nucl Med 48:1035–1046

31 Nakamoto Y, Chin BB, Cohade C, Osman M, Tatsumi M, Wahl RL (2004) PET/CT: artifacts caused by bowel motion. Nuclear Medicine Communications 25

32 Mostafapour S, Greuter M, van Snick JH, et al (2024) Ultra-low dose CT scanning for PET/CT. Med Phys 51:139–155

33 Shiri I, Salimi Y, Maghsudi M et al (2023) Differential privacy preserved federated transfer learning for multi-institutional (68)Ga-PET image artefact detection and disentanglement. Eur J Nucl Med Mol Imaging 51:40–53

34 D’Antonoli TA, Berger LK, Indrakanti AK et al (2024) TotalSegmentator MRI: Sequence-Independent Segmentation of 59 Anatomical Structures in MR images. arXiv preprint arXiv:240519492

35 Klyuzhin IS, Chaussé G, Bloise I et al (2024) PSMALJHornet: FullyLJautomated, multiLJtarget segmentation of healthy organs in PSMA PET/CT images. Medical Physics 51:1203–1216

36 Yazdani E, Karamzadeh-Ziarati N, Cheshmi SS et al (2024) Automated segmentation of lesions and organs at risk on [68Ga] Ga-PSMA-11 PET/CT images using self-supervised learning with Swin UNETR. Cancer Imaging 24:30

37 Wang X, Jemaa S, Fredrickson J et al (2022) Heart and bladder detection and segmentation on FDG PET/CT by deep learning. BMC Medical Imaging 22:58

38 Clement C, Xue S, Zhou X, et al (2024) Multi-Organ Segmentation on CT-free Total-Body Dynamic PET Scans. Soc Nuclear Med

39 Yushkevich PA, Piven J, Hazlett HC et al (2006) User-guided 3D active contour segmentation of anatomical structures: Significantly improved efficiency and reliability. Neuroimage 31:1116–1128

40 Isensee F, Jaeger PF, Kohl SAA, Petersen J, Maier-Hein KH (2021) nnU-Net: a self-configuring method for deep learning-based biomedical image segmentation. Nat Methods 18:203–211

41 Andrearczyk V, Oreiller V, Boughdad S et al (2023) Automatic Head and Neck Tumor segmentation and outcome prediction relying on FDG-PET/CT images: Findings from the second edition of the HECKTOR challenge. Med Image Anal 90:102972

42 Salimi Y, Mansouri Z, Hajianfar G, Sanaat A, Shiri I, Zaidi H (2024) Fully automated explainable abdominal CT contrast media phase classification using organ segmentation and machine learning. Med Phys. 10.1002/mp.17076

43 Shiri I, Arabi H, Salimi Y et al (2022) COLI-Net: Deep learning-assisted fully automated COVID-19 lung and infection pneumonia lesion detection and segmentation from chest computed tomography images. Int J Imaging Syst Technol 32:12–25

44 Akhavanallaf A, Fayad H, Salimi Y et al (2022) An update on computational anthropomorphic anatomical models. Digit Health 8:20552076221111941

45 Salimi Y, Akhavanallaf A, Mansouri Z, Shiri I, Zaidi H (2023) Real-time, acquisition parameter-free voxel-wise patient-specific Monte Carlo dose reconstruction in whole-body CT scanning using deep neural networks. Eur Radiol 33:9411–9424

46 Akramova R, Watanabe Y (2024) Radiomics as a measure superior to common similarity metrics for tumor segmentation performance evaluation. Journal of Applied Clinical Medical Physics n/a:e14442

47 Yazdani E, Karamzadeh-Ziarati N, Cheshmi SS et al (2024) Automated segmentation of lesions and organs at risk on [68Ga]Ga-PSMA-11 PET/CT images using self-supervised learning with Swin UNETR. Cancer Imaging 24:30

48 Klyuzhin IS, Chaussé G, Bloise I et al (2024) PSMA-Hornet: Fully-automated, multi-target segmentation of healthy organs in PSMA PET/CT images. Medical Physics 51:1203–1216

