## Supplementary material for "Deep Learning-powered CT-less Multi-tracer Organ Segmentation from PET Images: A solution for unreliable CT segmentation in PET/CT Imaging"

Table 1 summarizes Fivefold separated metrics for all tasks.

*Table 1: detailed performance of all tasks and organs separated by fold.*

| Task | Fold | Segment | Dice | Jaccard | Sensitivity | Specificity | Precision | Accuracy | Mean Surface Distance | Hausdorff Distance | Volume Difference | True Volume | Predicted Volume |
|---|---|---|---|---|---|---|---|---|---|---|---|---|---|
| 1 | fold_0 | Adrenal Glands | 0.567 ± 0.129 (0.07 to 0.787) | 0.406 ± 0.115 (0.036 to 0.649) | 0.589 ± 0.153 (0.092 to 0.979) | 1.0 ± 0.0 (1.0 to 1.0) | 0.58 ± 0.139 (0.056 to 0.874) | 1.0 ± 0.0 (1.0 to 1.0) | 2.378 ± 2.181 (0.683 to 13.86) | 10.981 ± 10.981 (2.721 to 60.476) | -0.133 ± 2.598 (-10.318 to 5.672) | 9.934 ± 4.226 (0.362 to 28.339) | 9.801 ± 3.777 (1.265 to 19.069) |
| | fold_0 | Aorta | 0.889 ± 0.031 (0.763 to 0.938) | 0.801 ± 0.049 (0.617 to 0.883) | 0.892 ± 0.046 (0.716 to 0.963) | 1.0 ± 0.0 (1.0 to 1.0) | 0.886 ± 0.037 (0.765 to 0.978) | 1.0 ± 0.0 (1.0 to 1.0) | 1.036 ± 0.566 (0.466 to 4.905) | 4.735 ± 7.513 (1.65 to 76.349) | 0.506 ± 23.956 (-165.515 to 56.93) | 300.789 ± 102.472 (127.99 to 736.853) | 301.294 ± 96.622 (147.978 to 571.339) |
| | fold_0 | Colon | 0.718 ± 0.088 (0.319 to 0.853) | 0.567 ± 0.1 (0.19 to 0.744) | 0.696 ± 0.097 (0.31 to 0.861) | 1.0 ± 0.0 (0.999 to 1.0) | 0.747 ± 0.096 (0.328 to 0.908) | 0.999 ± 0.0 (0.999 to 1.0) | 4.248 ± 2.338 (1.592 to 17.78) | 20.238 ± 16.877 (6.279 to 139.263) | -64.828 ± 117.26 (-416.673 to 201.476) | 938.693 ± 322.524 (206.423 to 2247.149) | 873.865 ± 304.734 (333.994 to 2065.863) |
| | fold_0 | Esophagues | 0.76 ± 0.076 (0.382 to 0.87) | 0.619 ± 0.092 (0.236 to 0.77) | 0.766 ± 0.1 (0.386 to 0.933) | 1.0 ± 0.0 (1.0 to 1.0) | 0.763 ± 0.079 (0.378 to 0.93) | 1.0 ± 0.0 (1.0 to 1.0) | 1.323 ± 1.014 (0.511 to 6.376) | 5.843 ± 6.124 (1.65 to 39.679) | -0.354 ± 8.556 (-50.468 to 13.644) | 48.682 ± 14.683 (27.21 to 118.221) | 48.327 ± 12.514 (23.856 to 83.67) |
| | fold_0 | Eyeballs | 0.762 ± 0.167 (0.033 to 0.937) | 0.639 ± 0.185 (0.017 to 0.882) | 0.778 ± 0.169 (0.124 to 0.975) | 1.0 ± 0.0 (1.0 to 1.0) | 0.75 ± 0.167 (0.019 to 0.947) | 1.0 ± 0.0 (1.0 to 1.0) | 1.7 ± 1.648 (0.32 to 11.345) | 4.853 ± 4.147 (1.4 to 36.23) | 1.003 ± 2.801 (-6.021 to 18.612) | 23.709 ± 3.508 (2.588 to 32.35) | 24.712 ± 2.476 (20.254 to 31.539) |
| | fold_0 | FemuralHeads | 0.909 ± 0.058 (0.572 to 0.95) | 0.837 ± 0.086 (0.401 to 0.905) | 0.922 ± 0.031 (0.809 to 0.97) | 1.0 ± 0.0 (0.999 to 1.0) | 0.9 ± 0.088 (0.412 to 0.966) | 1.0 ± 0.0 (0.999 to 1.0) | 2.881 ± 6.369 (0.277 to 38.306) | 36.439 ± 124.856 (1.65 to 816.813) | 35.742 ± 124.78 (-87.139 to 841.963) | 572.842 ± 253.123 (270.516 to 1238.422) | 608.584 ± 325.743 (273.751 to 1509.697) |
| | fold_0 | Gall Bladder | 0.582 ± 0.176 (0.035 to 0.895) | 0.431 ± 0.165 (0.018 to 0.81) | 0.594 ± 0.198 (0.023 to 0.961) | 1.0 ± 0.0 (1.0 to 1.0) | 0.619 ± 0.204 (0.065 to 1.0) | 1.0 ± 0.0 (1.0 to 1.0) | 5.045 ± 4.636 (1.216 to 34.005) | 23.084 ± 56.918 (3.625 to 534.276) | -3.005 ± 16.074 (-72.605 to 47.624) | 40.198 ± 27.087 (1.478 to 170.648) | 37.194 ± 23.031 (2.389 to 118.331) |
| | fold_0 | Heart | 0.916 ± 0.029 (0.808 to 0.957) | 0.846 ± 0.048 (0.678 to 0.918) | 0.926 ± 0.038 (0.809 to 0.987) | 1.0 ± 0.0 (1.0 to 1.0) | 0.909 ± 0.047 (0.784 to 0.983) | 1.0 ± 0.0 (0.999 to 1.0) | 2.824 ± 4.582 (0.747 to 35.073) | 20.199 ± 77.158 (2.333 to 628.647) | 11.188 ± 54.409 (-172.855 to 136.822) | 723.975 ± 202.822 (373.622 to 1423.395) | 735.163 ± 193.54 (413.619 to 1448.675) |
| | fold_0 | Iliacs | 0.889 ± 0.032 (0.745 to 0.934) | 0.801 ± 0.049 (0.594 to 0.876) | 0.883 ± 0.034 (0.723 to 0.933) | 1.0 ± 0.0 (1.0 to 1.0) | 0.896 ± 0.041 (0.745 to 0.958) | 1.0 ± 0.0 (0.999 to 1.0) | 0.806 ± 0.352 (0.242 to 2.566) | 3.197 ± 0.955 (1.65 to 7.966) | -11.097 ± 32.505 (-110.426 to 86.091) | 725.229 ± 139.067 (453.647 to 1074.451) | 714.132 ± 129.129 (441.206 to |

| | | | | | | | | | | | | | |
|---|---|---|---|---|---|---|---|---|---|---|---|---|---|
| | | | | | | | | | | | | | 1049.42) |
| | fold_0 | Kidneys | 0.851 ± 0.071 (0.456 to 0.935) | 0.747 ± 0.094 (0.295 to 0.879) | 0.856 ± 0.091 (0.321 to 0.967) | 1.0 ± 0.0 (1.0 to 1.0) | 0.854 ± 0.073 (0.511 to 0.942) | 1.0 ± 0.0 (0.999 to 1.0) | 2.041 ± 1.5 (0.62 to 11.448) | 11.498 ± 32.909 (3.25 to 332.901) | -3.783 ± 68.567 (-554.867 to 142.859) | 363.627 ± 95.857 (197.954 to 935.212) | 359.844 ± 73.315 (176.835 to 568.748) |
| | fold_0 | Liver | 0.915 ± 0.038 (0.667 to 0.963) | 0.846 ± 0.058 (0.5 to 0.929) | 0.93 ± 0.037 (0.84 to 0.991) | 1.0 ± 0.0 (0.998 to 1.0) | 0.905 ± 0.063 (0.507 to 0.984) | 1.0 ± 0.0 (0.998 to 1.0) | 2.486 ± 1.532 (0.965 to 12.91) | 10.59 ± 11.925 (3.25 to 80.996) | 46.103 ± 168.573 (-337.269 to 1088.435) | 1735.509 ± 470.312 (940.045 to 3537.851) | 1781.613 ± 461.593 (1048.559 to 3619.975) |
| | fold_0 | Lungs | 0.932 ± 0.016 (0.889 to 0.965) | 0.873 ± 0.028 (0.8 to 0.932) | 0.931 ± 0.031 (0.835 to 0.984) | 1.0 ± 0.0 (0.999 to 1.0) | 0.934 ± 0.027 (0.851 to 0.978) | 0.999 ± 0.0 (0.998 to 1.0) | 1.66 ± 0.589 (0.801 to 4.116) | 6.545 ± 3.123 (2.721 to 22.965) | -9.711 ± 183.065 (-516.2 to 413.155) | 3529.658 ± 993.168 (2091.873 to 7452.507) | 3519.947 ± 994.193 (2114.117 to 7115.582) |
| | fold_0 | Pancreas | 0.686 ± 0.118 (0.296 to 0.845) | 0.533 ± 0.126 (0.174 to 0.732) | 0.682 ± 0.14 (0.284 to 0.932) | 1.0 ± 0.0 (1.0 to 1.0) | 0.706 ± 0.123 (0.26 to 0.929) | 1.0 ± 0.0 (1.0 to 1.0) | 3.532 ± 3.817 (1.223 to 35.674) | 20.492 ± 82.353 (4.073 to 829.977) | -2.965 ± 19.151 (-75.681 to 34.582) | 98.071 ± 25.825 (26.524 to 207.465) | 95.106 ± 27.324 (31.632 to 179.929) |
| | fold_0 | Rectus Abdominus Muscle | 0.905 ± 0.027 (0.784 to 0.943) | 0.828 ± 0.043 (0.645 to 0.892) | 0.904 ± 0.033 (0.766 to 0.968) | 1.0 ± 0.0 (1.0 to 1.0) | 0.907 ± 0.037 (0.776 to 0.958) | 1.0 ± 0.0 (0.999 to 1.0) | 1.061 ± 0.36 (0.463 to 2.799) | 3.834 ± 1.037 (2.164 to 8.561) | -7.431 ± 58.13 (-150.784 to 120.623) | 1140.58 ± 229.552 (628.667 to 1802.654) | 1133.149 ± 204.94 (725.308 to 1651.869) |
| | fold_0 | RibCage | 0.668 ± 0.097 (0.353 to 0.836) | 0.51 ± 0.107 (0.214 to 0.719) | 0.672 ± 0.097 (0.338 to 0.847) | 1.0 ± 0.0 (0.999 to 1.0) | 0.669 ± 0.11 (0.37 to 0.903) | 0.999 ± 0.0 (0.998 to 1.0) | 1.93 ± 2.284 (0.439 to 15.203) | 12.235 ± 47.286 (2.164 to 353.094) | -0.519 ± 69.717 (-199.802 to 144.663) | 612.002 ± 134.132 (365.316 to 875.806) | 611.484 ± 105.057 (393.756 to 850.912) |
| | fold_0 | Sacrum | 0.877 ± 0.029 (0.783 to 0.918) | 0.782 ± 0.045 (0.644 to 0.849) | 0.888 ± 0.038 (0.724 to 0.95) | 1.0 ± 0.0 (1.0 to 1.0) | 0.867 ± 0.042 (0.74 to 0.94) | 1.0 ± 0.0 (1.0 to 1.0) | 1.149 ± 0.368 (0.588 to 2.477) | 4.714 ± 1.659 (2.721 to 12.772) | 5.458 ± 17.772 (-55.13 to 46.629) | 276.2 ± 47.026 (153.472 to 442.948) | 281.658 ± 40.252 (177.358 to 393.383) |
| | fold_0 | Spleen | 0.815 ± 0.169 (0.077 to 0.947) | 0.713 ± 0.18 (0.04 to 0.899) | 0.828 ± 0.187 (0.094 to 0.969) | 1.0 ± 0.0 (0.999 to 1.0) | 0.825 ± 0.148 (0.063 to 0.99) | 1.0 ± 0.0 (0.999 to 1.0) | 4.241 ± 9.122 (0.677 to 72.504) | 16.188 ± 32.419 (3.3 to 187.765) | -2.167 ± 75.939 (-423.248 to 393.035) | 281.318 ± 184.081 (17.979 to 1087.927) | 279.15 ± 178.281 (6.3 to 1106.21) |
| | fold_0 | Stomach | 0.748 ± 0.12 (0.098 to 0.894) | 0.61 ± 0.127 (0.051 to 0.808) | 0.755 ± 0.139 (0.058 to 0.931) | 1.0 ± 0.0 (1.0 to 1.0) | 0.756 ± 0.106 (0.3 to 0.954) | 1.0 ± 0.0 (0.999 to 1.0) | 3.564 ± 2.308 (1.277 to 19.377) | 13.051 ± 8.932 (3.983 to 65.645) | -3.801 ± 44.922 (-260.539 to 87.234) | 218.01 ± 75.547 (85.888 to 552.237) | 214.209 ± 67.862 (16.702 to 448.265) |
| | fold_0 | Urinary Bladder | 0.815 ± 0.125 (0.266 to 0.946) | 0.704 ± 0.151 (0.153 to 0.898) | 0.828 ± 0.128 (0.203 to 0.979) | 1.0 ± 0.0 (1.0 to 1.0) | 0.824 ± 0.158 (0.198 to 0.998) | 1.0 ± 0.0 (1.0 to 1.0) | 2.785 ± 2.926 (0.553 to 16.999) | 18.65 ± 64.578 (1.65 to 605.572) | -2.01 ± 28.185 (-112.665 to 77.591) | 116.369 ± 82.118 (29.568 to 570.094) | 114.359 ± 77.019 (43.212 to 542.495) |

| | | | | | | | | | | | | | |
|---|---|---|---|---|---|---|---|---|---|---|---|---|---|
| | fold_0 | Vertebrae | 0.857 ± 0.03 (0.699 to 0.905) | 0.751 ± 0.044 (0.537 to 0.826) | 0.859 ± 0.033 (0.697 to 0.917) | 1.0 ± 0.0 (0.999 to 1.0) | 0.857 ± 0.039 (0.701 to 0.932) | 1.0 ± 0.0 (0.999 to 1.0) | 1.171 ± 0.781 (0.507 to 5.808) | 4.358 ± 1.041 (2.721 to 8.731) | 0.26 ± 47.152 (-116.387 to 87.435) | 1036.684 ± 197.991 (654.494 to 1578.27) | 1036.944 ± 185.012 (681.416 to 1474.361) |
| | fold_0 | brain | 0.96 ± 0.028 (0.829 to 0.984) | 0.924 ± 0.049 (0.708 to 0.968) | 0.957 ± 0.029 (0.827 to 0.993) | 1.0 ± 0.0 (0.999 to 1.0) | 0.963 ± 0.035 (0.761 to 0.996) | 1.0 ± 0.0 (0.999 to 1.0) | 5.264 ± 10.699 (0.253 to 57.509) | 65.753 ± 235.816 (1.65 to 1613.038) | -7.971 ± 53.461 (-171.486 to 330.404) | 1434.458 ± 135.294 (1119.587 to 1793.715) | 1426.488 ± 131.954 (1119.487 to 1758.939) |
| | fold_0 | clavicle | 0.769 ± 0.069 (0.562 to 0.88) | 0.629 ± 0.088 (0.391 to 0.785) | 0.774 ± 0.074 (0.563 to 0.903) | 1.0 ± 0.0 (1.0 to 1.0) | 0.769 ± 0.088 (0.534 to 0.929) | 1.0 ± 0.0 (1.0 to 1.0) | 2.819 ± 6.034 (0.561 to 42.211) | 24.06 ± 78.898 (2.164 to 435.355) | -0.106 ± 10.025 (-21.706 to 21.081) | 86.222 ± 20.345 (46.629 to 140.16) | 86.116 ± 16.165 (57.519 to 120.615) |
| | fold_1 | Adrenal Glands | 0.563 ± 0.136 (0.127 to 0.763) | 0.403 ± 0.122 (0.068 to 0.616) | 0.567 ± 0.16 (0.091 to 0.892) | 1.0 ± 0.0 (1.0 to 1.0) | 0.591 ± 0.146 (0.118 to 0.846) | 1.0 ± 0.0 (1.0 to 1.0) | 2.21 ± 2.01 (0.728 to 12.133) | 9.914 ± 9.754 (3.0 to 62.806) | -0.479 ± 2.848 (-9.505 to 5.302) | 10.292 ± 4.905 (0.448 to 22.21) | 9.813 ± 4.63 (0.534 to 24.904) |
| | fold_1 | Aorta | 0.886 ± 0.055 (0.51 to 0.943) | 0.799 ± 0.075 (0.342 to 0.892) | 0.89 ± 0.047 (0.714 to 0.97) | 1.0 ± 0.0 (1.0 to 1.0) | 0.885 ± 0.07 (0.382 to 0.966) | 1.0 ± 0.0 (1.0 to 1.0) | 0.979 ± 0.438 (0.318 to 3.222) | 3.889 ± 1.84 (1.65 to 12.874) | -0.735 ± 21.211 (-128.063 to 36.713) | 298.408 ± 109.637 (30.406 to 764.945) | 297.673 ± 102.25 (61.16 to 636.883) |
| | fold_1 | Colon | 0.733 ± 0.091 (0.264 to 0.866) | 0.585 ± 0.103 (0.152 to 0.764) | 0.716 ± 0.09 (0.393 to 0.877) | 1.0 ± 0.0 (0.997 to 1.0) | 0.757 ± 0.098 (0.171 to 0.917) | 0.999 ± 0.0 (0.996 to 1.0) | 4.083 ± 2.806 (1.79 to 22.87) | 19.576 ± 18.522 (6.067 to 127.817) | -52.402 ± 172.889 (-665.442 to 1252.329) | 1011.259 ± 317.323 (440.609 to 2086.805) | 958.857 ± 300.353 (403.087 to 1914.125) |
| | fold_1 | Esophagues | 0.763 ± 0.081 (0.454 to 0.884) | 0.623 ± 0.098 (0.294 to 0.792) | 0.788 ± 0.081 (0.449 to 0.899) | 1.0 ± 0.0 (1.0 to 1.0) | 0.746 ± 0.099 (0.365 to 0.906) | 1.0 ± 0.0 (1.0 to 1.0) | 1.318 ± 1.05 (0.48 to 7.538) | 6.473 ± 8.644 (2.164 to 60.838) | 2.667 ± 8.315 (-43.775 to 40.673) | 45.76 ± 15.149 (18.711 to 127.411) | 48.428 ± 14.12 (15.029 to 85.362) |
| | fold_1 | Eyeballs | 0.745 ± 0.193 (0.035 to 0.946) | 0.625 ± 0.206 (0.018 to 0.898) | 0.77 ± 0.195 (0.034 to 0.98) | 1.0 ± 0.0 (1.0 to 1.0) | 0.729 ± 0.198 (0.035 to 0.966) | 1.0 ± 0.0 (1.0 to 1.0) | 1.976 ± 2.427 (0.254 to 17.222) | 5.631 ± 6.55 (1.5 to 52.738) | 1.17 ± 2.726 (-2.538 to 15.576) | 22.77 ± 3.309 (8.659 to 30.555) | 23.939 ± 2.114 (19.159 to 29.56) |
| | fold_1 | FemuralHeads | 0.912 ± 0.047 (0.747 to 0.953) | 0.841 ± 0.073 (0.596 to 0.91) | 0.922 ± 0.037 (0.77 to 0.97) | 1.0 ± 0.0 (0.999 to 1.0) | 0.904 ± 0.068 (0.668 to 0.968) | 1.0 ± 0.0 (0.999 to 1.0) | 3.715 ± 12.06 (0.274 to 91.175) | 31.33 ± 129.494 (2.027 to 953.635) | 22.329 ± 86.716 (-132.322 to 420.604) | 593.082 ± 318.076 (268.476 to 1650.921) | 615.411 ± 361.948 (267.829 to 1734.469) |
| | fold_1 | Gall Bladder | 0.629 ± 0.193 (0.067 to 0.876) | 0.485 ± 0.188 (0.035 to 0.779) | 0.644 ± 0.206 (0.125 to 1.0) | 1.0 ± 0.0 (1.0 to 1.0) | 0.664 ± 0.207 (0.035 to 1.0) | 1.0 ± 0.0 (1.0 to 1.0) | 4.271 ± 4.2 (1.125 to 32.885) | 18.895 ± 58.704 (3.585 to 561.782) | -1.077 ± 11.87 (-44.888 to 38.942) | 39.064 ± 20.171 (0.93 to 113.068) | 37.987 ± 20.105 (3.036 to 104.751) |
| | fold_1 | Heart | 0.915 ± 0.03 (0.777 to 0.958) | 0.844 ± 0.048 (0.636 to 0.92) | 0.928 ± 0.04 (0.789 to 0.984) | 1.0 ± 0.0 (1.0 to 1.0) | 0.904 ± 0.048 (0.716 to 0.988) | 1.0 ± 0.0 (0.999 to 1.0) | 2.259 ± 1.681 (0.874 to 14.806) | 9.2 ± 18.022 (2.721 to 130.154) | 18.132 ± 54.804 (-158.768 to 148.786) | 677.993 ± 180.741 (242.101 to 1278.433) | 696.125 ± 180.62 |

| | | | | | | | | | | | | | |
|---|---|---|---|---|---|---|---|---|---|---|---|---|---|
| | | | | | | | | | | | | | 8 (235.98 to 1306.798) |
| | fold_1 | Iliacs | 0.889 ± 0.033 (0.704 to 0.934) | 0.802 ± 0.051 (0.543 to 0.875) | 0.89 ± 0.034 (0.741 to 0.951) | 1.0 ± 0.0 (0.999 to 1.0) | 0.89 ± 0.043 (0.663 to 0.978) | 1.0 ± 0.0 (0.999 to 1.0) | 0.809 ± 0.335 (0.33 to 2.836) | 3.251 ± 1.003 (1.65 to 7.192) | -2.269 ± 34.526 (-120.594 to 98.737) | 738.883 ± 170.943 (433.108 to 1231.316) | 736.615 ± 157.452 (482.749 to 1230.558) |
| | fold_1 | Kidneys | 0.858 ± 0.09 (0.124 to 0.933) | 0.759 ± 0.101 (0.066 to 0.875) | 0.871 ± 0.102 (0.084 to 0.968) | 1.0 ± 0.0 (1.0 to 1.0) | 0.85 ± 0.091 (0.238 to 0.964) | 1.0 ± 0.0 (0.999 to 1.0) | 2.047 ± 3.596 (0.651 to 35.856) | 8.332 ± 12.266 (2.8 to 115.958) | 4.981 ± 43.76 (-262.807 to 100.175) | 351.091 ± 87.723 (85.693 to 611.929) | 356.072 ± 88.34 (142.275 to 596.917) |
| | fold_1 | Liver | 0.917 ± 0.072 (0.258 to 0.964) | 0.852 ± 0.083 (0.148 to 0.931) | 0.923 ± 0.088 (0.15 to 0.983) | 1.0 ± 0.0 (0.999 to 1.0) | 0.92 ± 0.043 (0.79 to 0.986) | 1.0 ± 0.001 (0.994 to 1.0) | 2.31 ± 2.192 (0.931 to 22.138) | 9.564 ± 9.941 (3.3 to 87.106) | -18.022 ± 315.341 (-2715.027 to 302.023) | 1720.799 ± 494.563 (832.3 to 4274.797) | 1702.777 ± 420.101 (527.691 to 3399.948) |
| | fold_1 | Lungs | 0.928 ± 0.02 (0.86 to 0.963) | 0.866 ± 0.034 (0.754 to 0.929) | 0.93 ± 0.034 (0.831 to 0.986) | 1.0 ± 0.0 (0.999 to 1.0) | 0.928 ± 0.033 (0.816 to 0.978) | 0.999 ± 0.0 (0.998 to 1.0) | 1.788 ± 0.624 (0.909 to 3.807) | 6.995 ± 3.133 (3.25 to 22.398) | 1.585 ± 202.341 (-633.814 to 489.69) | 3351.455 ± 972.968 (1419.115 to 6169.77) | 3353.039 ± 955.632 (1388.461 to 5945.136) |
| | fold_1 | Pancreas | 0.691 ± 0.108 (0.207 to 0.874) | 0.538 ± 0.117 (0.116 to 0.776) | 0.696 ± 0.131 (0.127 to 0.943) | 1.0 ± 0.0 (1.0 to 1.0) | 0.705 ± 0.115 (0.394 to 0.955) | 1.0 ± 0.0 (1.0 to 1.0) | 3.157 ± 1.789 (0.853 to 12.064) | 11.867 ± 10.058 (4.073 to 76.259) | -4.523 ± 23.177 (-145.957 to 39.015) | 96.624 ± 32.778 (24.384 to 215.428) | 92.102 ± 23.729 (22.011 to 135.503) |
| | fold_1 | Rectus Abdominus Muscle | 0.904 ± 0.02 (0.844 to 0.943) | 0.826 ± 0.033 (0.73 to 0.892) | 0.908 ± 0.023 (0.842 to 0.949) | 1.0 ± 0.0 (1.0 to 1.0) | 0.902 ± 0.033 (0.798 to 0.963) | 1.0 ± 0.0 (0.999 to 1.0) | 1.077 ± 0.302 (0.613 to 2.475) | 4.233 ± 3.419 (2.164 to 32.72) | 1.323 ± 51.695 (-171.53 to 141.528) | 1116.229 ± 266.088 (470.467 to 1811.2) | 1117.552 ± 240.237 (469.273 to 1661.463) |
| | fold_1 | RibCage | 0.658 ± 0.105 (0.306 to 0.838) | 0.499 ± 0.111 (0.18 to 0.721) | 0.676 ± 0.105 (0.317 to 0.845) | 1.0 ± 0.0 (0.999 to 1.0) | 0.644 ± 0.116 (0.295 to 0.855) | 0.999 ± 0.0 (0.998 to 1.0) | 1.757 ± 1.823 (0.5 to 13.934) | 8.321 ± 28.771 (2.164 to 286.368) | 25.919 ± 61.817 (-219.261 to 168.952) | 596.569 ± 144.874 (350.018 to 967.922) | 622.488 ± 120.048 (386.366 to 917.291) |
| | fold_1 | Sacrum | 0.873 ± 0.035 (0.733 to 0.919) | 0.776 ± 0.053 (0.578 to 0.851) | 0.886 ± 0.038 (0.737 to 0.958) | 1.0 ± 0.0 (1.0 to 1.0) | 0.86 ± 0.047 (0.687 to 0.943) | 1.0 ± 0.0 (1.0 to 1.0) | 1.371 ± 1.31 (0.566 to 13.145) | 6.69 ± 16.534 (3.25 to 165.742) | 7.864 ± 16.252 (-36.19 to 52.153) | 279.815 ± 55.789 (188.704 to 435.17) | 287.679 ± 53.641 (199.503 to 425.531) |
| | fold_1 | Spleen | 0.824 ± 0.146 (0.026 to 0.945) | 0.72 ± 0.151 (0.013 to 0.896) | 0.833 ± 0.147 (0.125 to 0.989) | 1.0 ± 0.0 (1.0 to 1.0) | 0.825 ± 0.159 (0.015 to 0.984) | 1.0 ± 0.0 (0.999 to 1.0) | 3.21 ± 5.519 (0.842 to 44.592) | 12.641 ± 22.72 (3.3 to 145.0) | 3.359 ± 37.262 (-141.727 to 164.783) | 249.878 ± 149.856 (2.558 to 992.042) | 253.237 ± 147.885 (21.733 to 1003.09) |

| | | | | | | | | | | | | |
|---|---|---|---|---|---|---|---|---|---|---|---|---|
| | fold_1 | Stomach | 0.781 ± 0.066 (0.472 to 0.875) | 0.646 ± 0.082 (0.309 to 0.777) | 0.786 ± 0.096 (0.317 to 0.953) | 1.0 ± 0.0 (1.0 to 1.0) | 0.787 ± 0.073 (0.538 to 0.926) | 1.0 ± 0.0 (1.0 to 1.0) | 3.04 ± 1.651 (1.082 to 15.761) | 12.07 ± 9.899 (4.073 to 79.05) | -0.85 ± 29.774 (-118.843 to 52.785) | 209.776 ± 70.123 (38.618 to 410.003) | 208.926 ± 67.551 (33.232 to 437.008) |
| | fold_1 | Urinary Bladder | 0.84 ± 0.089 (0.358 to 0.952) | 0.733 ± 0.117 (0.218 to 0.909) | 0.846 ± 0.127 (0.293 to 0.999) | 1.0 ± 0.0 (1.0 to 1.0) | 0.854 ± 0.112 (0.46 to 0.999) | 1.0 ± 0.0 (1.0 to 1.0) | 2.205 ± 1.582 (0.575 to 8.607) | 9.475 ± 10.02 (2.164 to 58.313) | -1.426 ± 28.82 (-137.992 to 124.057) | 129.547 ± 101.597 (39.22 to 543.222) | 128.121 ± 103.748 (41.092 to 550.189) |
| | fold_1 | Vertebrae | 0.86 ± 0.026 (0.733 to 0.903) | 0.755 ± 0.039 (0.579 to 0.822) | 0.865 ± 0.033 (0.735 to 0.925) | 1.0 ± 0.0 (0.999 to 1.0) | 0.855 ± 0.033 (0.714 to 0.911) | 1.0 ± 0.0 (0.999 to 1.0) | 1.162 ± 0.893 (0.58 to 6.935) | 4.396 ± 1.21 (2.721 to 10.0) | 8.684 ± 55.635 (-286.598 to 111.73) | 1047.11 ± 248.367 (630.508 to 1841.677) | 1055.794 ± 231.178 (615.579 to 1854.664) |
| | fold_1 | brain | 0.956 ± 0.037 (0.774 to 0.986) | 0.918 ± 0.061 (0.631 to 0.972) | 0.955 ± 0.032 (0.821 to 0.988) | 1.0 ± 0.0 (0.999 to 1.0) | 0.959 ± 0.051 (0.638 to 0.992) | 1.0 ± 0.0 (0.999 to 1.0) | 6.743 ± 13.915 (0.17 to 79.015) | 72.447 ± 247.18 (1.65 to 1586.1) | -7.022 ± 77.774 (-207.217 to 466.326) | 1427.277 ± 184.528 (834.192 to 1876.296) | 1420.256 ± 166.762 (1029.886 to 1901.825) |
| | fold_1 | clavicle | 0.765 ± 0.078 (0.475 to 0.887) | 0.626 ± 0.098 (0.312 to 0.797) | 0.769 ± 0.084 (0.428 to 0.908) | 1.0 ± 0.0 (1.0 to 1.0) | 0.765 ± 0.088 (0.508 to 0.9) | 1.0 ± 0.0 (1.0 to 1.0) | 3.048 ± 5.854 (0.416 to 30.502) | 28.692 ± 84.95 (2.164 to 420.52) | -0.233 ± 8.303 (-28.365 to 20.827) | 83.877 ± 20.869 (46.479 to 133.82) | 83.644 ± 17.639 (51.506 to 126.184) |
| | fold_2 | Adrenal Glands | 0.582 ± 0.122 (0.263 to 0.788) | 0.421 ± 0.118 (0.152 to 0.65) | 0.586 ± 0.139 (0.268 to 0.938) | 1.0 ± 0.0 (1.0 to 1.0) | 0.592 ± 0.132 (0.232 to 0.825) | 1.0 ± 0.0 (1.0 to 1.0) | 1.989 ± 1.574 (0.587 to 12.324) | 9.371 ± 9.637 (2.776 to 81.272) | -0.386 ± 2.383 (-13.536 to 4.745) | 10.461 ± 4.933 (0.796 to 29.993) | 10.076 ± 4.279 (1.543 to 23.677) |
| | fold_2 | Aorta | 0.89 ± 0.039 (0.682 to 0.94) | 0.803 ± 0.059 (0.518 to 0.887) | 0.894 ± 0.04 (0.702 to 0.964) | 1.0 ± 0.0 (1.0 to 1.0) | 0.886 ± 0.047 (0.663 to 0.963) | 1.0 ± 0.0 (1.0 to 1.0) | 0.942 ± 0.39 (0.42 to 2.8) | 3.631 ± 1.717 (1.65 to 13.612) | 2.307 ± 13.498 (-37.97 to 33.491) | 284.599 ± 85.132 (124.571 to 519.199) | 286.906 ± 84.079 (119.997 to 515.361) |
| | fold_2 | Colon | 0.744 ± 0.07 (0.517 to 0.872) | 0.598 ± 0.086 (0.348 to 0.773) | 0.738 ± 0.085 (0.413 to 0.891) | 1.0 ± 0.0 (0.999 to 1.0) | 0.756 ± 0.07 (0.586 to 0.884) | 0.999 ± 0.0 (0.996 to 1.0) | 3.636 ± 1.475 (1.475 to 10.764) | 15.851 ± 9.084 (5.59 to 49.578) | -36.544 ± 134.888 (-861.967 to 266.983) | 995.688 ± 374.174 (398.405 to 2239.721) | 959.144 ± 322.41 (400.848 to 1977.665) |
| | fold_2 | Esophagues | 0.767 ± 0.074 (0.489 to 0.868) | 0.627 ± 0.091 (0.324 to 0.767) | 0.778 ± 0.084 (0.434 to 0.931) | 1.0 ± 0.0 (1.0 to 1.0) | 0.76 ± 0.085 (0.451 to 0.904) | 1.0 ± 0.0 (1.0 to 1.0) | 1.141 ± 0.474 (0.455 to 2.791) | 4.611 ± 2.642 (2.027 to 21.417) | 1.058 ± 5.51 (-16.82 to 14.758) | 46.686 ± 11.344 (26.922 to 87.809) | 47.744 ± 11.174 (26.723 to 81.372) |
| | fold_2 | Eyeballs | 0.798 ± 0.142 (0.296 to 0.944) | 0.683 ± 0.167 (0.174 to 0.894) | 0.815 ± 0.147 (0.31 to 0.98) | 1.0 ± 0.0 (1.0 to 1.0) | 0.784 ± 0.143 (0.283 to 0.956) | 1.0 ± 0.0 (1.0 to 1.0) | 1.355 ± 1.223 (0.223 to 5.697) | 4.057 ± 2.422 (1.5 to 12.0) | 0.909 ± 1.762 (-2.861 to 7.615) | 23.87 ± 2.504 (16.332 to 29.913) | 24.779 ± 2.392 (20.362 to 31.539) |
| | fold_2 | FemuralHeads | 0.9 ± 0.059 (0.625 to 0.951) | 0.823 ± 0.089 (0.455 to 0.906) | 0.908 ± 0.058 (0.618 to 0.978) | 1.0 ± 0.0 (0.999 to 1.0) | 0.895 ± 0.077 (0.623 to 0.966) | 1.0 ± 0.0 (0.999 to 1.0) | 2.846 ± 5.983 (0.336 to 41.95) | 21.254 ± 82.509 (1.65 to 758.864) | 20.607 ± 91.511 (-168.5 to 365.913) | 646.684 ± 357.994 (278.727 to 2037.978) | 667.291 ± |

| | | | | | | | | | | | | | |
|---|---|---|---|---|---|---|---|---|---|---|---|---|---|
| | | | | | | | | | | | | | 388.219 (262.753 to 1869.478) |
| | fold_2 | Gall Bladder | 0.564 ± 0.211 (0.033 to 0.868) | 0.421 ± 0.196 (0.017 to 0.766) | 0.58 ± 0.228 (0.018 to 1.0) | 1.0 ± 0.0 (1.0 to 1.0) | 0.603 ± 0.225 (0.062 to 0.982) | 1.0 ± 0.0 (1.0 to 1.0) | 5.074 ± 4.117 (0.712 to 23.081) | 16.049 ± 17.821 (3.69 to 150.685) | -2.953 ± 13.906 (-46.728 to 35.77) | 36.563 ± 23.633 (1.543 to 124.187) | 33.61 ± 19.845 (0.697 to 106.427) |
| | fold_2 | Heart | 0.916 ± 0.027 (0.819 to 0.956) | 0.846 ± 0.045 (0.694 to 0.916) | 0.918 ± 0.045 (0.789 to 0.986) | 1.0 ± 0.0 (1.0 to 1.0) | 0.917 ± 0.045 (0.771 to 0.991) | 1.0 ± 0.0 (0.999 to 1.0) | 3.268 ± 5.921 (0.856 to 48.244) | 26.745 ± 94.118 (3.424 to 575.119) | -0.849 ± 57.84 (-148.794 to 147.5) | 707.506 ± 176.402 (430.167 to 1171.852) | 706.657 ± 173.117 (443.188 to 1235.589) |
| | fold_2 | Iliacs | 0.884 ± 0.04 (0.649 to 0.933) | 0.794 ± 0.059 (0.48 to 0.874) | 0.893 ± 0.042 (0.643 to 0.954) | 1.0 ± 0.0 (1.0 to 1.0) | 0.877 ± 0.048 (0.655 to 0.958) | 1.0 ± 0.0 (0.999 to 1.0) | 0.852 ± 0.41 (0.367 to 3.23) | 3.254 ± 1.029 (1.65 to 7.823) | 11.879 ± 34.749 (-106.544 to 115.552) | 717.462 ± 142.7 (463.254 to 1057.344) | 729.341 ± 136.769 (460.685 to 1070.521) |
| | fold_2 | Kidneys | 0.852 ± 0.064 (0.603 to 0.939) | 0.746 ± 0.09 (0.432 to 0.885) | 0.876 ± 0.069 (0.565 to 0.978) | 1.0 ± 0.0 (1.0 to 1.0) | 0.836 ± 0.091 (0.475 to 0.968) | 1.0 ± 0.0 (1.0 to 1.0) | 2.593 ± 4.481 (0.856 to 38.752) | 13.367 ± 31.957 (2.8 to 286.833) | 13.377 ± 43.478 (-134.064 to 146.405) | 343.865 ± 100.799 (105.483 to 617.661) | 357.242 ± 89.79 (151.481 to 583.727) |
| | fold_2 | Liver | 0.918 ± 0.031 (0.723 to 0.96) | 0.851 ± 0.05 (0.566 to 0.923) | 0.919 ± 0.043 (0.789 to 0.987) | 1.0 ± 0.0 (0.999 to 1.0) | 0.921 ± 0.051 (0.598 to 0.991) | 1.0 ± 0.0 (0.998 to 1.0) | 2.371 ± 1.45 (0.939 to 13.292) | 9.589 ± 11.296 (3.625 to 101.985) | -9.717 ± 155.95 (-449.776 to 782.984) | 1737.755 ± 508.133 (805.862 to 3928.898) | 1728.038 ± 471.22 (921.118 to 3479.122) |
| | fold_2 | Lungs | 0.929 ± 0.021 (0.846 to 0.962) | 0.868 ± 0.035 (0.734 to 0.927) | 0.936 ± 0.031 (0.834 to 0.99) | 1.0 ± 0.0 (0.999 to 1.0) | 0.924 ± 0.037 (0.809 to 0.985) | 0.999 ± 0.0 (0.998 to 1.0) | 1.785 ± 0.889 (0.885 to 8.091) | 7.102 ± 3.522 (3.25 to 22.362) | 41.534 ± 206.316 (-630.549 to 508.229) | 3410.902 ± 1011.871 (1690.079 to 7746.793) | 3452.436 ± 998.038 (1656.29 to 7392.072) |
| | fold_2 | Pancreas | 0.689 ± 0.11 (0.258 to 0.851) | 0.535 ± 0.119 (0.148 to 0.74) | 0.687 ± 0.124 (0.261 to 0.909) | 1.0 ± 0.0 (1.0 to 1.0) | 0.701 ± 0.118 (0.256 to 0.906) | 1.0 ± 0.0 (1.0 to 1.0) | 3.456 ± 2.923 (1.23 to 21.84) | 15.355 ± 45.81 (4.073 to 465.848) | -1.744 ± 14.76 (-34.243 to 31.035) | 98.198 ± 27.556 (37.364 to 180.318) | 96.454 ± 28.29 (26.829 to 196.833) |
| | fold_2 | Rectus Abdominus Muscle | 0.903 ± 0.026 (0.796 to 0.938) | 0.824 ± 0.042 (0.66 to 0.884) | 0.897 ± 0.028 (0.793 to 0.942) | 1.0 ± 0.0 (1.0 to 1.0) | 0.909 ± 0.037 (0.729 to 0.965) | 1.0 ± 0.0 (0.999 to 1.0) | 1.1 ± 0.429 (0.507 to 4.085) | 4.3 ± 4.616 (2.164 to 49.154) | -19.087 ± 53.634 (-195.126 to 110.77) | 1135.653 ± 285.06 (551.04 to 1917.325) | 1116.566 ± 258.13 (655.29 to 1732.704) |
| | fold_2 | RibCage | 0.658 ± 0.105 (0.341 to 0.825) | 0.499 ± 0.111 (0.205 to 0.702) | 0.676 ± 0.113 (0.323 to 0.912) | 1.0 ± 0.0 (0.999 to 1.0) | 0.645 ± 0.111 (0.338 to 0.864) | 0.999 ± 0.0 (0.998 to 1.0) | 1.69 ± 1.525 (0.516 to 12.453) | 5.924 ± 3.943 (2.164 to 25.625) | 21.033 ± 66.758 (-171.835 to 175.592) | 588.875 ± 148.729 (312.616 to 1013.432) | 609.908 ± 119.5 (365.714 to 958.081) |
| | fold_2 | Sacrum | 0.874 ± 0.036 (0.723 to 0.918) | 0.778 ± 0.054 (0.566 to 0.848) | 0.895 ± 0.04 (0.712 to 0.952) | 1.0 ± 0.0 (1.0 to 1.0) | 0.856 ± 0.046 (0.69 to 0.924) | 1.0 ± 0.0 (1.0 to 1.0) | 1.21 ± 0.48 (0.656 to 3.186) | 4.988 ± 2.747 (2.8 to 20.364) | 12.577 ± 17.506 (-38.569 to 77.632) | 271.754 ± 47.333 (174.82 to 440.51) | 284.331 ± 50.28 |

| | | | | | | | | | | | | | |
|---|---|---|---|---|---|---|---|---|---|---|---|---|---|
| | | | | | | | | | | | | | (197.214 to 422.097) |
| | fold_2 | Spleen | 0.824 ± 0.136 (0.073 to 0.953) | 0.717 ± 0.148 (0.038 to 0.91) | 0.84 ± 0.118 (0.049 to 0.967) | 1.0 ± 0.0 (0.999 to 1.0) | 0.829 ± 0.139 (0.039 to 0.975) | 1.0 ± 0.0 (0.999 to 1.0) | 3.269 ± 4.921 (0.629 to 35.809) | 12.992 ± 22.846 (2.774 to 145.831) | 3.421 ± 62.621 (-328.64 to 290.467) | 253.513 ± 145.702 (24.413 to 893.522) | 256.934 ± 142.596 (17.815 to 856.665) |
| | fold_2 | Stomach | 0.775 ± 0.09 (0.274 to 0.897) | 0.64 ± 0.107 (0.158 to 0.814) | 0.785 ± 0.101 (0.465 to 0.949) | 1.0 ± 0.0 (1.0 to 1.0) | 0.776 ± 0.109 (0.194 to 0.945) | 1.0 ± 0.0 (0.999 to 1.0) | 3.199 ± 1.809 (1.081 to 16.302) | 12.582 ± 9.482 (3.645 to 60.701) | 0.539 ± 46.057 (-159.21 to 231.004) | 233.658 ± 79.649 (61.612 to 504.904) | 234.196 ± 71.074 (98.483 to 444.69) |
| | fold_2 | Urinary Bladder | 0.824 ± 0.107 (0.307 to 0.957) | 0.713 ± 0.138 (0.181 to 0.917) | 0.819 ± 0.144 (0.185 to 0.979) | 1.0 ± 0.0 (1.0 to 1.0) | 0.852 ± 0.111 (0.515 to 1.0) | 1.0 ± 0.0 (0.999 to 1.0) | 3.309 ± 6.451 (0.686 to 60.699) | 21.101 ± 86.794 (2.164 to 863.773) | -11.881 ± 53.119 (-358.544 to 36.925) | 127.986 ± 109.567 (35.508 to 820.058) | 116.105 ± 82.27 (47.125 to 482.361) |
| | fold_2 | Vertebrae | 0.857 ± 0.032 (0.746 to 0.898) | 0.752 ± 0.047 (0.595 to 0.816) | 0.871 ± 0.033 (0.75 to 0.947) | 1.0 ± 0.0 (0.999 to 1.0) | 0.846 ± 0.042 (0.687 to 0.906) | 1.0 ± 0.0 (0.999 to 1.0) | 1.057 ± 0.374 (0.417 to 2.766) | 4.442 ± 1.87 (2.8 to 19.242) | 29.631 ± 49.663 (-78.328 to 210.591) | 1015.636 ± 206.177 (637.773 to 1660.42) | 1045.267 ± 207.835 (671.129 to 1686.447) |
| | fold_2 | brain | 0.965 ± 0.019 (0.894 to 0.984) | 0.933 ± 0.034 (0.809 to 0.969) | 0.963 ± 0.024 (0.828 to 0.994) | 1.0 ± 0.0 (1.0 to 1.0) | 0.967 ± 0.023 (0.839 to 0.993) | 1.0 ± 0.0 (0.999 to 1.0) | 7.295 ± 14.731 (0.22 to 68.121) | 83.024 ± 265.349 (1.65 to 1634.946) | -6.232 ± 46.657 (-278.08 to 184.812) | 1433.353 ± 160.777 (1157.667 to 1958.28) | 1427.121 ± 146.051 (1170.847 to 1918.846) |
| | fold_2 | clavicle | 0.76 ± 0.081 (0.517 to 0.897) | 0.62 ± 0.1 (0.348 to 0.814) | 0.772 ± 0.09 (0.502 to 0.935) | 1.0 ± 0.0 (1.0 to 1.0) | 0.752 ± 0.091 (0.526 to 0.929) | 1.0 ± 0.0 (1.0 to 1.0) | 2.379 ± 3.933 (0.585 to 31.3) | 17.895 ± 56.586 (2.164 to 335.413) | 1.563 ± 8.952 (-24.527 to 24.605) | 84.593 ± 20.569 (41.304 to 138.841) | 86.157 ± 17.577 (44.58 to 129.734) |
| | fold_3 | Adrenal Glands | 0.554 ± 0.135 (0.069 to 0.812) | 0.395 ± 0.123 (0.036 to 0.683) | 0.575 ± 0.143 (0.137 to 1.0) | 1.0 ± 0.0 (1.0 to 1.0) | 0.567 ± 0.158 (0.036 to 0.844) | 1.0 ± 0.0 (1.0 to 1.0) | 2.233 ± 1.914 (0.441 to 13.342) | 10.832 ± 9.138 (3.3 to 56.416) | -0.319 ± 2.562 (-8.833 to 5.081) | 10.533 ± 4.264 (0.1 to 23.102) | 10.214 ± 3.696 (2.787 to 22.171) |
| | fold_3 | Aorta | 0.879 ± 0.043 (0.75 to 0.944) | 0.787 ± 0.066 (0.6 to 0.894) | 0.881 ± 0.052 (0.68 to 0.954) | 1.0 ± 0.0 (1.0 to 1.0) | 0.881 ± 0.055 (0.702 to 0.98) | 1.0 ± 0.0 (1.0 to 1.0) | 1.025 ± 0.478 (0.44 to 2.777) | 4.279 ± 2.36 (1.65 to 19.347) | -2.849 ± 26.142 (-148.197 to 56.731) | 296.749 ± 93.302 (124.161 to 565.466) | 293.9 ± 81.893 (153.322 to 510.611) |
| | fold_3 | Colon | 0.722 ± 0.079 (0.483 to 0.851) | 0.57 ± 0.094 (0.318 to 0.741) | 0.707 ± 0.093 (0.4 to 0.856) | 1.0 ± 0.0 (0.999 to 1.0) | 0.743 ± 0.086 (0.512 to 0.881) | 0.999 ± 0.0 (0.997 to 1.0) | 4.01 ± 1.689 (1.674 to 13.156) | 18.786 ± 11.18 (6.0 to 79.351) | -57.855 ± 144.082 (-552.279 to 254.139) | 1055.41 ± 467.908 (302.675 to 3643.973) | 997.554 ± 423.98 (338.747 to 3275.996) |
| | fold_3 | Esophagues | 0.736 ± 0.089 (0.492 to 0.874) | 0.59 ± 0.107 (0.326 to 0.776) | 0.737 ± 0.105 (0.417 to 0.903) | 1.0 ± 0.0 (1.0 to 1.0) | 0.741 ± 0.092 (0.516 to 0.898) | 1.0 ± 0.0 (1.0 to 1.0) | 1.348 ± 0.735 (0.487 to 4.554) | 6.004 ± 3.731 (2.164 to 20.55) | -0.851 ± 8.361 (-58.106 to 16.065) | 49.456 ± 14.47 (23.041 to 127.956) | 48.604 ± 11.904 (27.32 to |

| | | | | | | | | | | | | | |
|---|---|---|---|---|---|---|---|---|---|---|---|---|---|
| | | | | | | | | | | | | | 83.651 ) |
| | fold _3 | Eyeballs | 0.744 ± 0.169 (0.125 to 0.934) | 0.617 ± 0.193 (0.067 to 0.877) | 0.765 ± 0.166 (0.123 to 0.977) | 1.0 ± 0.0 (1.0 to 1.0) | 0.727 ± 0.175 (0.126 to 0.956) | 1.0 ± 0.0 (1.0 to 1.0) | 1.859 ± 1.88 (0.233 to 12.674) | 5.579 ± 5.745 (1.65 to 53.2) | 1.228 ± 2.305 (-2.198 to 11.534) | 23.293 ± 2.905 (10.646 to 32.23) | 24.521 ± 2.395 (20.055 to 30.359) |
| | fold _3 | FemuralHeads | 0.906 ± 0.05 (0.737 to 0.952) | 0.831 ± 0.078 (0.583 to 0.909) | 0.91 ± 0.046 (0.731 to 0.968) | 1.0 ± 0.0 (0.999 to 1.0) | 0.903 ± 0.067 (0.61 to 0.986) | 1.0 ± 0.0 (0.999 to 1.0) | 3.764 ± 9.932 (0.3 to 65.281) | 43.317 ± 153.801 (1.65 to 878.918) | 14.475 ± 72.862 (-176.711 to 397.083) | 610.356 ± 318.94 (290.082 to 1754.452) | 624.831 ± 347.241 (288.378 to 1904.122) |
| | fold _3 | Gall Bladder | 0.585 ± 0.185 (0.065 to 0.876) | 0.435 ± 0.174 (0.034 to 0.779) | 0.578 ± 0.195 (0.078 to 0.965) | 1.0 ± 0.0 (1.0 to 1.0) | 0.632 ± 0.218 (0.052 to 0.984) | 1.0 ± 0.0 (1.0 to 1.0) | 4.668 ± 3.237 (1.098 to 18.446) | 17.478 ± 15.783 (3.585 to 93.582) | -5.058 ± 14.091 (-64.36 to 28.266) | 41.595 ± 25.183 (1.923 to 119.682) | 36.536 ± 19.261 (4.063 to 101.618) |
| | fold _3 | Heart | 0.907 ± 0.043 (0.642 to 0.951) | 0.833 ± 0.065 (0.473 to 0.907) | 0.909 ± 0.062 (0.515 to 0.983) | 1.0 ± 0.0 (1.0 to 1.0) | 0.909 ± 0.049 (0.733 to 0.986) | 1.0 ± 0.0 (0.999 to 1.0) | 3.418 ± 6.644 (0.768 to 54.011) | 19.801 ± 55.243 (3.3 to 414.074) | -4.131 ± 84.389 (-579.455 to 183.031) | 742.623 ± 211.533 (369.25 to 1616.578) | 738.492 ± 195.463 (341.259 to 1528.795) |
| | fold _3 | Iliacs | 0.887 ± 0.025 (0.794 to 0.93) | 0.797 ± 0.04 (0.658 to 0.869) | 0.881 ± 0.029 (0.803 to 0.948) | 1.0 ± 0.0 (1.0 to 1.0) | 0.893 ± 0.035 (0.77 to 0.963) | 1.0 ± 0.0 (0.999 to 1.0) | 0.762 ± 0.269 (0.292 to 1.488) | 3.374 ± 0.804 (1.65 to 5.76) | -12.033 ± 31.949 (-89.774 to 92.421) | 733.033 ± 152.179 (406.869 to 1168.554) | 721.0 ± 139.874 (462.903 to 1128.793) |
| | fold _3 | Kidneys | 0.861 ± 0.047 (0.674 to 0.919) | 0.758 ± 0.069 (0.508 to 0.85) | 0.866 ± 0.073 (0.612 to 0.98) | 1.0 ± 0.0 (1.0 to 1.0) | 0.861 ± 0.059 (0.695 to 0.979) | 1.0 ± 0.0 (1.0 to 1.0) | 1.731 ± 0.844 (0.631 to 5.717) | 7.738 ± 5.612 (3.0 to 46.961) | 0.049 ± 41.602 (-136.677 to 106.655) | 362.699 ± 86.239 (181.24 to 615.279) | 362.749 ± 78.342 (185.967 to 610.412) |
| | fold _3 | Liver | 0.915 ± 0.034 (0.74 to 0.965) | 0.845 ± 0.055 (0.587 to 0.933) | 0.92 ± 0.05 (0.614 to 0.989) | 1.0 ± 0.0 (0.998 to 1.0) | 0.914 ± 0.045 (0.78 to 0.983) | 1.0 ± 0.0 (0.998 to 1.0) | 2.556 ± 2.125 (0.95 to 20.632) | 10.362 ± 14.911 (3.585 to 153.515) | 13.908 ± 183.633 (-1080.497 to 858.388) | 1846.155 ± 465.836 (927.2 to 3655.59) | 1860.063 ± 497.034 (926.155 to 4513.978) |
| | fold _3 | Lungs | 0.921 ± 0.025 (0.836 to 0.96) | 0.855 ± 0.042 (0.718 to 0.924) | 0.92 ± 0.036 (0.824 to 0.991) | 1.0 ± 0.0 (0.998 to 1.0) | 0.925 ± 0.045 (0.745 to 0.987) | 0.999 ± 0.0 (0.998 to 1.0) | 1.92 ± 0.801 (0.74 to 5.56) | 8.693 ± 6.673 (3.3 to 61.279) | -22.913 ± 243.963 (-561.237 to 821.999) | 3391.335 ± 1048.515 (1598.013 to 7779.98) | 3368.422 ± 1007.233 (1625.83 to 7871.742) |
| | fold _3 | Pancreas | 0.686 ± 0.136 (0.035 to 0.863) | 0.536 ± 0.136 (0.018 to 0.76) | 0.699 ± 0.138 (0.067 to 0.923) | 1.0 ± 0.0 (1.0 to 1.0) | 0.685 ± 0.149 (0.024 to 0.909) | 1.0 ± 0.0 (1.0 to 1.0) | 3.548 ± 3.437 (1.049 to 26.231) | 12.998 ± 15.232 (3.69 to 124.326) | 0.483 ± 17.157 (-72.152 to 49.515) | 97.48 ± 29.338 (18.114 to 176.076) | 97.963 ± 22.284 (50.112 to 149.883) |
| | fold _3 | Rectus Abdominus Muscle | 0.905 ± 0.022 (0.836 to 0.941) | 0.827 ± 0.035 (0.719 to 0.889) | 0.901 ± 0.027 (0.803 to 0.954) | 1.0 ± 0.0 (1.0 to 1.0) | 0.91 ± 0.033 (0.784 to 0.963) | 1.0 ± 0.0 (0.999 to 1.0) | 1.062 ± 0.366 (0.536 to 2.94) | 4.392 ± 4.009 (2.164 to 44.054) | -15.02 ± 57.854 (-144.116 to 184.527) | 1203.822 ± 242.748 (772.882 to 1837.978) | 1188.802 ± 220.393 (752.0 |

| | | | | | | | | | | | | | |
|---|---|---|---|---|---|---|---|---|---|---|---|---|---|
| | | | | | | | | | | | | | 31 to 1710.93) |
| | fold_3 | RibCage | 0.635 ± 0.121 (0.322 to 0.834) | 0.476 ± 0.123 (0.192 to 0.715) | 0.635 ± 0.125 (0.293 to 0.867) | 1.0 ± 0.0 (0.999 to 1.0) | 0.638 ± 0.126 (0.294 to 0.846) | 0.999 ± 0.0 (0.998 to 1.0) | 1.818 ± 1.986 (0.413 to 17.943) | 9.389 ± 36.351 (2.5 to 368.479) | -8.644 ± 70.106 (-338.519 to 137.062) | 605.082 ± 145.935 (345.73 to 1098.437) | 596.438 ± 113.541 (363.624 to 841.277) |
| | fold_3 | Sacrum | 0.87 ± 0.03 (0.778 to 0.914) | 0.771 ± 0.046 (0.637 to 0.841) | 0.886 ± 0.03 (0.815 to 0.951) | 1.0 ± 0.0 (1.0 to 1.0) | 0.856 ± 0.044 (0.699 to 0.944) | 1.0 ± 0.0 (1.0 to 1.0) | 1.214 ± 0.535 (0.501 to 4.785) | 4.961 ± 1.6 (3.25 to 12.218) | 9.469 ± 15.701 (-39.769 to 67.579) | 273.886 ± 50.025 (153.571 to 414.533) | 283.355 ± 51.466 (180.692 to 433.891) |
| | fold_3 | Spleen | 0.827 ± 0.113 (0.037 to 0.939) | 0.717 ± 0.128 (0.019 to 0.885) | 0.85 ± 0.116 (0.032 to 0.985) | 1.0 ± 0.0 (0.999 to 1.0) | 0.815 ± 0.136 (0.045 to 0.983) | 1.0 ± 0.0 (0.999 to 1.0) | 3.349 ± 6.699 (0.769 to 65.125) | 12.239 ± 17.513 (3.645 to 133.66) | 13.523 ± 46.116 (-89.742 to 166.616) | 291.433 ± 170.706 (53.795 to 1318.596) | 304.956 ± 179.915 (60.364 to 1358.887) |
| | fold_3 | Stomach | 0.751 ± 0.086 (0.431 to 0.887) | 0.608 ± 0.105 (0.275 to 0.797) | 0.744 ± 0.112 (0.32 to 0.934) | 1.0 ± 0.0 (1.0 to 1.0) | 0.769 ± 0.086 (0.525 to 0.946) | 1.0 ± 0.0 (0.999 to 1.0) | 3.971 ± 4.452 (1.064 to 45.52) | 19.097 ± 47.295 (4.073 to 451.7) | -10.052 ± 40.639 (-169.246 to 67.977) | 235.504 ± 75.131 (120.08 to 490.277) | 225.452 ± 67.553 (107.988 to 473.567) |
| | fold_3 | Urinary Bladder | 0.811 ± 0.129 (0.377 to 0.957) | 0.699 ± 0.159 (0.232 to 0.918) | 0.833 ± 0.153 (0.258 to 1.0) | 1.0 ± 0.0 (1.0 to 1.0) | 0.82 ± 0.155 (0.286 to 0.995) | 1.0 ± 0.0 (1.0 to 1.0) | 6.473 ± 31.048 (0.415 to 314.872) | 38.089 ± 154.103 (2.721 to 1071.159) | 0.286 ± 46.653 (-271.171 to 263.98) | 110.787 ± 62.508 (29.211 to 437.854) | 111.073 ± 63.037 (40.267 to 418.481) |
| | fold_3 | Vertebrae | 0.854 ± 0.026 (0.786 to 0.904) | 0.746 ± 0.039 (0.648 to 0.824) | 0.859 ± 0.034 (0.783 to 0.937) | 1.0 ± 0.0 (0.999 to 1.0) | 0.85 ± 0.033 (0.754 to 0.944) | 1.0 ± 0.0 (0.999 to 1.0) | 1.971 ± 5.629 (0.397 to 44.3) | 27.296 ± 159.1 (2.5 to 1191.428) | 9.09 ± 51.628 (-165.962 to 119.35) | 1045.295 ± 201.76 (620.754 to 1582.341) | 1054.385 ± 196.749 (647.129 to 1556.264) |
| | fold_3 | brain | 0.957 ± 0.029 (0.817 to 0.985) | 0.918 ± 0.05 (0.691 to 0.971) | 0.954 ± 0.028 (0.811 to 0.987) | 1.0 ± 0.0 (0.999 to 1.0) | 0.96 ± 0.039 (0.734 to 0.995) | 1.0 ± 0.0 (0.999 to 1.0) | 5.319 ± 10.73 (0.222 to 58.101) | 45.61 ± 129.627 (1.65 to 524.97) | -11.703 ± 56.796 (-186.49 to 316.107) | 1433.118 ± 171.702 (989.942 to 1830.68) | 1421.415 ± 149.003 (1120.35 to 1779.74) |
| | fold_3 | clavicle | 0.758 ± 0.098 (0.243 to 0.889) | 0.619 ± 0.11 (0.139 to 0.801) | 0.761 ± 0.109 (0.242 to 0.915) | 1.0 ± 0.0 (1.0 to 1.0) | 0.76 ± 0.101 (0.245 to 0.931) | 1.0 ± 0.0 (1.0 to 1.0) | 2.316 ± 4.254 (0.458 to 30.063) | 19.899 ± 65.693 (2.164 to 433.222) | -0.796 ± 10.469 (-39.396 to 19.109) | 85.199 ± 19.545 (47.226 to 147.948) | 84.404 ± 15.87 (54.143 to 124.509) |
| | fold_4 | Adrenal Glands | 0.539 ± 0.151 (0.115 to 0.769) | 0.383 ± 0.131 (0.061 to 0.624) | 0.542 ± 0.182 (0.084 to 0.869) | 1.0 ± 0.0 (1.0 to 1.0) | 0.562 ± 0.149 (0.132 to 0.818) | 1.0 ± 0.0 (1.0 to 1.0) | 2.346 ± 2.267 (0.715 to 17.73) | 10.172 ± 8.203 (2.721 to 56.885) | -0.238 ± 2.524 (-5.889 to 6.777) | 9.378 ± 3.675 (1.883 to 19.328) | 9.14 ± 4.332 (0.581 to 19.955) |
| | fold_4 | Aorta | 0.879 ± 0.054 (0.553 to 0.941) | 0.787 ± 0.078 (0.382 to 0.888) | 0.882 ± 0.066 (0.412 to 0.965) | 1.0 ± 0.0 (1.0 to 1.0) | 0.879 ± 0.054 (0.716 to 0.969) | 1.0 ± 0.0 (0.999 to 1.0) | 1.132 ± 1.041 (0.437 to 10.144) | 5.219 ± 8.812 (1.65 to 83.028) | -1.795 ± 38.685 (-338.991 to 47.762) | 298.416 ± 103.09 (108.289 to 664.297) | 296.62 ± 93.645 (126.668 to |

| | | | | | | | | | | | | | |
|---|---|---|---|---|---|---|---|---|---|---|---|---|---|
| | | | | | | | | | | | | | 580.099) |
| | fold_4 | Colon | 0.707 ± 0.117 (0.22 to 0.845) | 0.558 ± 0.122 (0.124 to 0.732) | 0.699 ± 0.126 (0.171 to 0.9) | 1.0 ± 0.0 (0.998 to 1.0) | 0.724 ± 0.118 (0.156 to 0.889) | 0.999 ± 0.0 (0.997 to 1.0) | 4.344 ± 3.257 (2.071 to 29.569) | 19.738 ± 20.117 (7.169 to 160.171) | -28.267 ± 132.297 (-323.067 to 656.325) | 959.659 ± 388.079 (249.068 to 2951.187) | 931.392 ± 395.236 (139.09 to 3187.553) |
| | fold_4 | Esophagues | 0.745 ± 0.096 (0.359 to 0.88) | 0.602 ± 0.111 (0.219 to 0.785) | 0.749 ± 0.099 (0.435 to 0.9) | 1.0 ± 0.0 (1.0 to 1.0) | 0.747 ± 0.109 (0.306 to 0.917) | 1.0 ± 0.0 (1.0 to 1.0) | 1.343 ± 0.88 (0.495 to 6.062) | 6.293 ± 5.908 (1.65 to 37.079) | 0.286 ± 6.266 (-17.571 to 23.826) | 47.256 ± 11.891 (23.715 to 73.485) | 47.542 ± 12.015 (22.443 to 80.583) |
| | fold_4 | Eyeballs | 0.74 ± 0.174 (0.132 to 0.936) | 0.614 ± 0.197 (0.071 to 0.88) | 0.767 ± 0.17 (0.129 to 0.971) | 1.0 ± 0.0 (1.0 to 1.0) | 0.724 ± 0.188 (0.135 to 0.979) | 1.0 ± 0.0 (1.0 to 1.0) | 2.497 ± 4.039 (0.195 to 28.452) | 14.383 ± 58.138 (1.4 to 469.786) | 1.377 ± 3.744 (-6.668 to 18.562) | 23.125 ± 3.869 (7.219 to 34.934) | 24.502 ± 3.024 (18.371 to 42.05) |
| | fold_4 | FemuralHeads | 0.9 ± 0.071 (0.57 to 0.949) | 0.825 ± 0.101 (0.399 to 0.902) | 0.908 ± 0.053 (0.697 to 0.961) | 1.0 ± 0.0 (0.999 to 1.0) | 0.897 ± 0.092 (0.43 to 0.974) | 1.0 ± 0.0 (0.999 to 1.0) | 4.792 ± 12.184 (0.349 to 64.14) | 62.472 ± 180.276 (2.164 to 927.371) | 20.525 ± 114.607 (-248.272 to 679.426) | 551.925 ± 240.552 (283.892 to 1796.425) | 572.45 ± 287.803 (296.714 to 1809.413) |
| | fold_4 | Gall Bladder | 0.574 ± 0.191 (0.025 to 0.883) | 0.426 ± 0.178 (0.012 to 0.791) | 0.589 ± 0.216 (0.017 to 1.0) | 1.0 ± 0.0 (1.0 to 1.0) | 0.607 ± 0.209 (0.042 to 0.944) | 1.0 ± 0.0 (1.0 to 1.0) | 6.594 ± 17.987 (0.573 to 166.286) | 32.592 ± 100.324 (3.0 to 654.46) | -1.73 ± 14.334 (-40.673 to 72.572) | 38.194 ± 28.713 (6.396 to 181.298) | 36.464 ± 25.836 (0.484 to 144.44) |
| | fold_4 | Heart | 0.911 ± 0.036 (0.68 to 0.952) | 0.838 ± 0.056 (0.515 to 0.908) | 0.91 ± 0.052 (0.66 to 0.979) | 1.0 ± 0.0 (0.999 to 1.0) | 0.914 ± 0.048 (0.701 to 0.988) | 1.0 ± 0.0 (0.999 to 1.0) | 3.377 ± 7.52 (0.779 to 65.114) | 23.342 ± 82.678 (3.0 to 690.28) | -6.569 ± 62.282 (-177.798 to 170.012) | 713.31 ± 207.486 (395.573 to 1340.887) | 706.741 ± 195.068 (383.679 to 1339.543) |
| | fold_4 | Iliacs | 0.886 ± 0.048 (0.497 to 0.931) | 0.798 ± 0.063 (0.331 to 0.871) | 0.882 ± 0.051 (0.5 to 0.952) | 1.0 ± 0.0 (0.999 to 1.0) | 0.891 ± 0.052 (0.494 to 0.95) | 1.0 ± 0.0 (0.999 to 1.0) | 0.818 ± 0.52 (0.369 to 4.999) | 3.386 ± 1.601 (1.65 to 16.291) | -9.45 ± 33.299 (-97.677 to 91.034) | 740.995 ± 140.794 (513.42 to 1045.092) | 731.546 ± 127.52 (511.921 to 1008.139) |
| | fold_4 | Kidneys | 0.834 ± 0.099 (0.113 to 0.939) | 0.725 ± 0.112 (0.06 to 0.886) | 0.862 ± 0.083 (0.49 to 1.0) | 1.0 ± 0.0 (1.0 to 1.0) | 0.82 ± 0.11 (0.06 to 0.972) | 1.0 ± 0.0 (0.999 to 1.0) | 2.435 ± 3.274 (0.651 to 31.86) | 12.007 ± 22.089 (2.721 to 189.391) | 12.227 ± 42.642 (-104.233 to 233.05) | 331.425 ± 104.345 (14.872 to 682.262) | 343.653 ± 92.475 (130.008 to 699.53) |
| | fold_4 | Liver | 0.907 ± 0.059 (0.506 to 0.955) | 0.834 ± 0.081 (0.338 to 0.914) | 0.908 ± 0.083 (0.361 to 0.99) | 1.0 ± 0.0 (0.999 to 1.0) | 0.911 ± 0.052 (0.77 to 0.985) | 0.999 ± 0.0 (0.996 to 1.0) | 2.672 ± 2.256 (1.065 to 20.145) | 10.33 ± 11.031 (3.3 to 93.208) | -14.825 ± 249.194 (-1859.814 to 490.879) | 1677.562 ± 460.644 (944.567 to 3242.969) | 1662.737 ± 451.086 (971.612 to 3353.585) |
| | fold_4 | Lungs | 0.927 ± 0.025 (0.788 to 0.966) | 0.865 ± 0.042 (0.65 to 0.934) | 0.93 ± 0.034 (0.834 to 0.985) | 1.0 ± 0.0 (0.998 to 1.0) | 0.926 ± 0.042 (0.738 to 0.984) | 0.999 ± 0.0 (0.998 to 1.0) | 1.969 ± 1.145 (0.712 to 8.668) | 8.534 ± 7.19 (2.8 to 57.037) | 14.728 ± 223.816 (-460.723 to 725.012) | 3464.008 ± 786.221 (2027.958 to 6074.63) | 3478.736 ± 777.095 (2021.407 to |

| | | | | | | | | | | | | | |
|---|---|---|---|---|---|---|---|---|---|---|---|---|---|
| | | | | | | | | | | | | | 6246.274) |
| | fold_4 | Pancreas | 0.657 ± 0.149 (0.136 to 0.859) | 0.505 ± 0.15 (0.073 to 0.752) | 0.674 ± 0.174 (0.081 to 0.898) | 1.0 ± 0.0 (1.0 to 1.0) | 0.659 ± 0.139 (0.228 to 0.868) | 1.0 ± 0.0 (1.0 to 1.0) | 4.111 ± 3.381 (1.128 to 17.977) | 18.754 ± 40.257 (4.073 to 377.423) | 0.689 ± 21.845 (-105.979 to 46.931) | 91.111 ± 30.058 (26.822 to 211.346) | 91.8 ± 27.641 (4.985 to 180.642) |
| | fold_4 | Rectus Abdominus Muscle | 0.899 ± 0.031 (0.792 to 0.94) | 0.818 ± 0.049 (0.655 to 0.887) | 0.899 ± 0.034 (0.772 to 0.96) | 1.0 ± 0.0 (1.0 to 1.0) | 0.9 ± 0.037 (0.789 to 0.961) | 1.0 ± 0.0 (0.999 to 1.0) | 1.114 ± 0.457 (0.503 to 3.585) | 4.111 ± 1.756 (2.164 to 16.291) | -5.759 ± 47.975 (-168.989 to 101.12) | 1131.374 ± 266.285 (637.683 to 2062.282) | 1125.615 ± 244.94 (702.665 to 1907.91) |
| | fold_4 | RibCage | 0.638 ± 0.12 (0.141 to 0.802) | 0.479 ± 0.122 (0.076 to 0.669) | 0.651 ± 0.124 (0.146 to 0.876) | 1.0 ± 0.0 (0.999 to 1.0) | 0.63 ± 0.129 (0.137 to 0.859) | 0.999 ± 0.0 (0.998 to 1.0) | 1.451 ± 0.722 (0.486 to 5.769) | 5.774 ± 2.831 (2.333 to 20.35) | 16.732 ± 66.539 (-167.744 to 154.644) | 597.371 ± 129.881 (380.917 to 870.686) | 614.103 ± 112.365 (370.492 to 872.723) |
| | fold_4 | Sacrum | 0.869 ± 0.066 (0.306 to 0.926) | 0.773 ± 0.076 (0.181 to 0.863) | 0.883 ± 0.072 (0.296 to 0.96) | 1.0 ± 0.0 (1.0 to 1.0) | 0.857 ± 0.068 (0.317 to 0.942) | 1.0 ± 0.0 (0.999 to 1.0) | 1.24 ± 0.843 (0.544 to 7.641) | 5.124 ± 4.023 (2.721 to 36.549) | 7.612 ± 15.685 (-38.428 to 42.797) | 270.81 ± 43.398 (175.794 to 392.874) | 278.422 ± 40.414 (181.816 to 411.002) |
| | fold_4 | Spleen | 0.802 ± 0.134 (0.235 to 0.931) | 0.687 ± 0.155 (0.133 to 0.87) | 0.818 ± 0.159 (0.2 to 0.99) | 1.0 ± 0.0 (0.999 to 1.0) | 0.808 ± 0.128 (0.26 to 0.983) | 1.0 ± 0.0 (0.999 to 1.0) | 3.835 ± 5.506 (0.788 to 40.373) | 15.07 ± 24.224 (3.3 to 148.105) | 4.371 ± 62.923 (-218.365 to 358.258) | 258.401 ± 152.285 (87.833 to 961.443) | 262.772 ± 165.303 (30.397 to 1161.505) |
| | fold_4 | Stomach | 0.737 ± 0.118 (0.165 to 0.905) | 0.595 ± 0.129 (0.09 to 0.827) | 0.745 ± 0.128 (0.209 to 0.917) | 1.0 ± 0.0 (1.0 to 1.0) | 0.745 ± 0.14 (0.137 to 0.942) | 1.0 ± 0.0 (0.999 to 1.0) | 4.39 ± 5.1 (1.046 to 47.861) | 18.112 ± 22.778 (3.3 to 181.97) | -4.645 ± 53.965 (-332.877 to 125.089) | 223.433 ± 93.848 (32.325 to 642.2) | 218.788 ± 75.906 (80.418 to 456.221) |
| | fold_4 | Urinary Bladder | 0.817 ± 0.101 (0.356 to 0.956) | 0.702 ± 0.132 (0.216 to 0.916) | 0.833 ± 0.128 (0.349 to 1.0) | 1.0 ± 0.0 (1.0 to 1.0) | 0.825 ± 0.138 (0.363 to 0.998) | 1.0 ± 0.0 (0.999 to 1.0) | 3.921 ± 12.485 (0.288 to 123.936) | 21.531 ± 79.797 (2.164 to 770.266) | -6.115 ± 56.303 (-454.91 to 147.699) | 130.339 ± 144.811 (32.695 to 1087.055) | 124.224 ± 107.962 (44.027 to 632.145) |
| | fold_4 | Vertebrae | 0.852 ± 0.039 (0.671 to 0.909) | 0.745 ± 0.057 (0.505 to 0.833) | 0.856 ± 0.041 (0.698 to 0.927) | 1.0 ± 0.0 (0.999 to 1.0) | 0.85 ± 0.049 (0.621 to 0.952) | 0.999 ± 0.0 (0.998 to 1.0) | 1.125 ± 0.892 (0.505 to 8.804) | 4.472 ± 1.198 (2.721 to 9.883) | 7.249 ± 59.314 (-142.498 to 203.325) | 1055.719 ± 189.039 (685.048 to 1494.508) | 1062.968 ± 187.816 (716.044 to 1584.46) |
| | fold_4 | brain | 0.95 ± 0.04 (0.74 to 0.986) | 0.908 ± 0.065 (0.587 to 0.972) | 0.951 ± 0.043 (0.674 to 0.989) | 1.0 ± 0.0 (0.999 to 1.0) | 0.952 ± 0.048 (0.651 to 0.997) | 1.0 ± 0.0 (0.999 to 1.0) | 8.574 ± 19.458 (0.225 to 145.333) | 77.978 ± 216.849 (1.65 to 1531.881) | -4.063 ± 80.139 (-312.245 to 450.666) | 1443.833 ± 169.025 (874.037 to 1877.639) | 1439.77 ± 143.457 (1104.11 to 1782.082) |
| | fold_4 | clavicle | 0.757 ± 0.089 (0.395 to 0.89) | 0.617 ± 0.106 (0.246 to 0.802) | 0.757 ± 0.095 (0.388 to 0.931) | 1.0 ± 0.0 (1.0 to 1.0) | 0.761 ± 0.099 (0.403 to 0.932) | 1.0 ± 0.0 (1.0 to 1.0) | 2.097 ± 2.506 (0.392 to 16.747) | 15.534 ± 49.595 (2.164 to 301.846) | -0.651 ± 8.807 (-32.546 to 18.006) | 85.755 ± 18.844 (50.83 to 129.081) | 85.104 ± 17.225 (51.23 to |

| | | | | | | | | | | | | | |
|---|---|---|---|---|---|---|---|---|---|---|---|---|---|
| | | | | | | | | | | | | | 123.97<br>1) |

Figure 1 compares the box plot dice for all tasks.

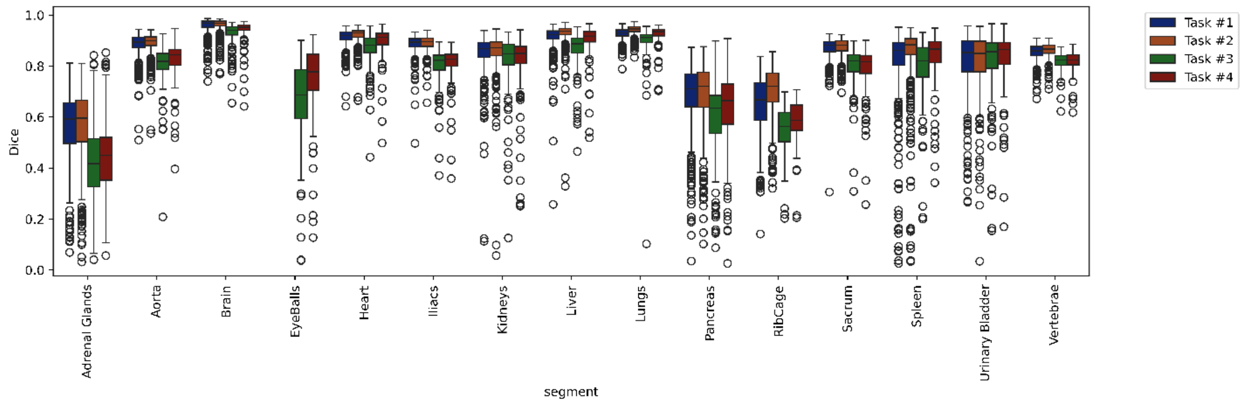


*Figure 1: the performance for all tasks compared*

Supplementary Figure 2 shows an example of a dynamic FDG image and the segmentation s generated by task #2 model.

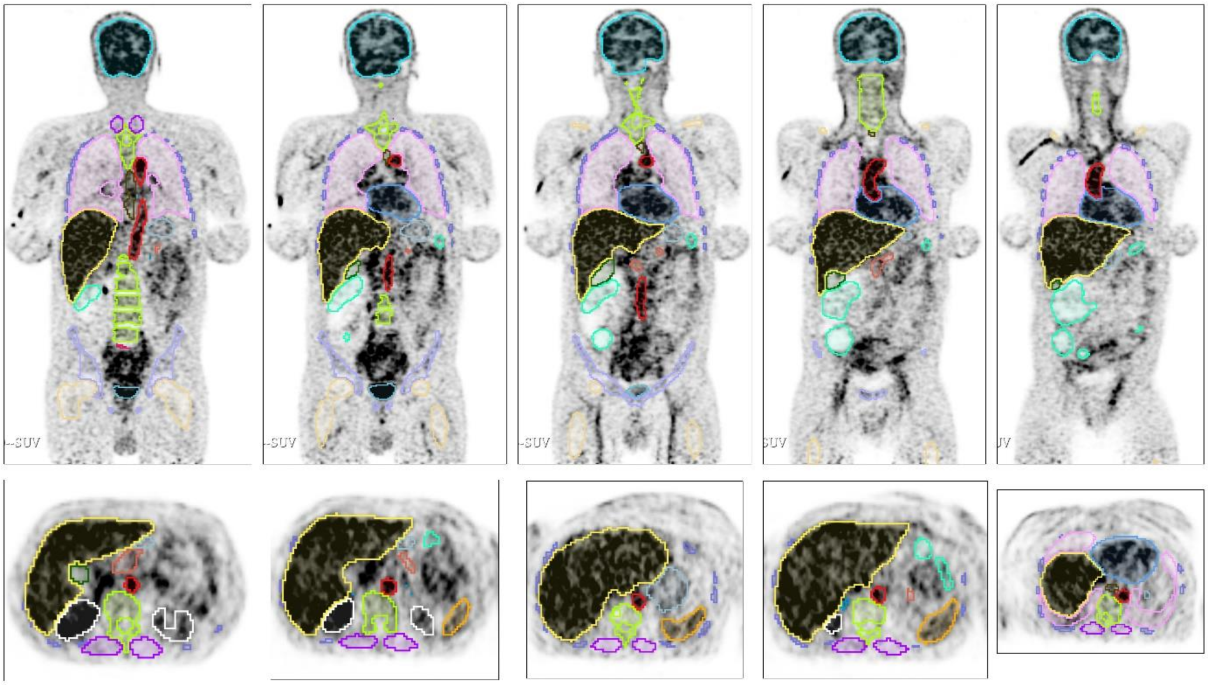


*Figure 2: example of a dynamic noisy frame segmented by task #2 model. The voxel spacing in the SUV image is 4 mm isotropic, the segmentation mask is resampled to image dimensions leading to the pixelated appearance of the segmentations.*
